# Assessing Statistical Practices of Existing Artificial Intelligence (AI) Models for Lung Cancer Detection, Prognosis, and Risk Prediction: A Cross-Sectional Meta-Research Study Supplemented by Human and Large Language Model (LLM)-Directed Quality Appraisal

**DOI:** 10.64898/2025.12.23.25342849

**Authors:** Yiren Hou, Timothy Ward, Cheng-Han Yang, Ebony Jernigan, Giorgio Caturegli, Daniel Boffa, Bhramar Mukherjee

## Abstract

Artificial intelligence (AI) models with medical images as input data are increasingly proposed to support clinical decisions in lung cancer screening. To assess how these models are developed, evaluated, and reported, and to identify gaps in best ***statistical practices,*** we conducted a cross-sectional meta-research study of OpenAlex-indexed studies (January 1, 2023, to June 30, 2025) that developed image-based AI tools to detect lung cancer, predict prognosis, or estimate future risk. Thirty-six studies met our inclusion criteria. Study quality and reporting were appraised using three approaches: subjective ratings from two statisticians and two clinicians, scoring from two AI agents (GPT-5 and Gemini 2.5 Pro), and a guideline-based checklist from the Critical Appraisal and Data Extraction for Systematic Reviews of Prediction Modelling Studies (CHARMS). Convolutional neural networks were used in most of the included studies (69%). Area under the curve was the most frequently reported metric (81%). Our meta-research study also highlights common lapses in these 36 studies, including limited external test set use (39%), insufficient subgroup analyses (28%), and a substantial lack of adherence to established prediction-model reporting guidelines. AI-based quality scoring aligned better with CHARMS-based scores than did human scoring. Spearman correlations with CHARMS were weaker for statisticians/clinicians (p ≤ 0.46) than for the two AI agents (GPT-5 p = 0.66; Gemini 2.5 Pro p = 0.56). Overall, future research should prioritize standardized reporting, use of external test sets, and model performance assessment across subpopulations. Large language models (LLMs) offer a supportive role in providing guideline-driven appraisals to complement human judgment in evaluating AI-based prediction models.

**1-2 Sentence Description:** This cross-sectional meta-research study synthesizes recent studies that developed artificial intelligence (AI)-driven predictive models using medical images to detect lung cancer, predict prognosis, or estimate future risk, highlighting methodological trends, limitations in model testing and subgroup analyses, and advocating for the need for greater transparency, reliability, quality assessment, and adherence to established reporting guidelines in such studies. Quality assessment of the models carried out by LLMs, human statisticians and clinicians indicates chatbots are more aligned with recommended guidelines than humans.

## Introduction

### The rise of artificial intelligence-driven models in predicting lung cancer outcomes

The public release of the Chat Generative Pre-trained Transformer (ChatGPT) [1] in November 2022 marked a transformative milestone in the advancement of generative artificial intelligence (GenAI), initiating great interest in GenAI-related models, e.g., generative adversarial networks (GANs), variational autoencoders (VAEs), and transformer-based models, across disciplines [2]. Although tumor phenotype classification has traditionally been conducted at the discretion of trained pathologists, early AI applications in oncology have largely relied on convolutional neural networks (CNNs) to characterize tumor phenotypes [3]. The extent to which newer GenAI architectures have been adopted in cancer imaging prediction models remains unclear [4]. Unlike the traditional regression approach, the shift toward deep learning and GenAI tools introduces methodological opacity [5], as modern deep learning architectures such as CNNs are often defined by millions of parameters and complex design choices [6,7]. To fully understand the reliability of AI models designed to support decision-making in oncology, we need to look beyond reported prediction metrics by objectively evaluating the model design, architecture, and generalizability.

Lung cancer remains the leading cause of cancer-related mortality worldwide, accounting for an estimated 1.8 million deaths annually [8]. As a result, improving the care of lung cancer patients is a global priority. Imaging modalities such as computed tomography (CT) play a central role in the management of lung cancer patients, making this a particularly attractive patient population for AI-based decision support tools [9]. Recently, investigators have applied AI methodologies to address largely three types of decision making: (1) detecting the presence of cancer (detection) [10,11], (2) predicting survival or cancer recurrence (prognosis) [12–14], and (3) predicting future cancers (risk prediction) [15,16]. While the era of lung cancer radiomics and AI is relatively young, there is already considerable optimism for major improvements in early diagnosis and prevention in this population [16,17].

### Generalizability of AI-based lung cancer risk prediction models: the case of the Sybil model

A notable example of AI innovation in lung cancer is Sybil [16], a deep learning model that predicts 1- to 6-year lung cancer risk from a single low-dose CT (LDCT) scan. While the model was trained using radiologist-annotated data, it operates in practice without requiring annotations or clinical variables, using just the images. In its original development, Sybil was trained on 28,162 LDCTs from the National Lung Screening Trial (NLST) [18] and tested on three independent cohorts: a held-out NLST set (6,282 scans), a U.S. screening program at Massachusetts General Hospital (8,821 scans), and a diverse dataset from Chang Gung Memorial Hospital in Taiwan (CGMH) (12,280 scans). Across these cohorts, Sybil demonstrated robust performance, achieving 1-year AUCs of 0.92, 0.86, and 0.94, respectively. Perhaps most surprisingly, Sybil retained substantial predictive ability even among scans that radiologists judged to be unconcerning. More specifically, in the subset of scans interpreted as radiologically negative (Lung-RADS 1-2), Sybil achieved a 1-year AUC of 0.86.

Despite the large validation cohorts of Sybil, recent studies suggest there is room to improve the generalizability of the model. A recent analysis by Li et al. [19] observed that performance dropped when applied to a prospective cohort of patients presented with persistent pulmonary nodules. In this prospective cohort, the 1-year AUC of Sybil was 0.67 (95% CI 0.60-0.74). Calibration analysis suggested that Sybil underestimated patient cancer risk for low-risk nodules and overestimated cancer risk for high-risk nodules. This finding indicates that AI-based predictive models of imaging studies may perform differently depending on the context in which the scan was obtained (e.g., screening versus surveillance of an established nodule).

### Reporting frameworks for AI model development and objectives of this cross-sectional meta-research study

One of the more disconcerting elements of AI-based decision support tools has been the inability to delineate which CT scan features contribute most to a patient’s risk determination. This opacity can make it harder for clinicians to trust these models and make it challenging to vet the models for compliance with best practices. One approach to ensuring transparency in studies developing clinical prediction models has been the establishment of reporting guidelines [20–27] and critical appraisal tools. The Critical Appraisal and Data Extraction for Systematic Reviews of Prediction Modelling Studies (CHARMS) [28] and Prediction model study Risk of Bias Assessment Tool (PROBAST) [29] are two commonly used methodological frameworks that guide how individual prediction model studies are extracted and appraised, particularly when they are extracted in systematic reviews. While both checklists cover similar domains, PROBAST formally rates risk of bias and applicability across four domains (participants, predictors, outcome, and analysis), whereas CHARMS structures data extraction and methodological appraisal into a set of key items that prediction model studies should report.

Recently, the Standards for Reporting Diagnostic Accuracy-AI statement (STARD-AI), published in September 2025 as the successor to the 2015 STARD guidelines, introduced a minimum set of criteria for reporting AI-centered diagnostic accuracy studies and addressed issues unique to AI, particularly in algorithmic and data practices [30]. These tools encourage reproducibility and comparability across studies. However, in the lung cancer imaging literature, the degree to which studies adhere to these standards in practice and how such adherence reveals concrete modeling choices (architecture, statistical parameters, and design decisions) remain unclear.

This work has two objectives. First and primarily, we conduct a cross-sectional meta-research study of literature published from January 1, 2023, to June 30, 2025, that developed novel AI-based prediction models using medical imaging to address key outcome endpoints—namely, detection, prognosis, and risk prediction of lung cancer. We summarize trends in study design, choice of methodology and algorithms, uncertainty quantification, evaluation practices, and assess the field’s adherence to standardized reporting guidelines. We focus on critically examining the underlying model architectures, statistical parameters, and design choices of these prediction tools. Second, in an exploratory analysis nested within this meta-research study, we evaluate whether general-purpose GenAI tools can assist with study appraisal. We achieve this by comparing quality scores for the included studies provided by GPT-5 [31] and Gemini 2.5 Pro [32] with CHARMS-based scores and subjective ratings from two statisticians and two clinicians. These GenAI tools are used only to appraise the reporting and quality of published studies (not to build a new image-based lung cancer prediction tool). We then quantify the agreement among these different scoring approaches using correlation analyses.

## Methods

### Search strategy and screening procedure

Studies published between January 1, 2023, and March 27, 2025, inclusive, were identified through the OpenAlex database [33] using the web application in the first search, with a second supplementary search designed to capture additional studies published between March 27, 2025, and June 30, 2025, inclusive. We searched for studies that developed and applied AI methods, including machine learning (ML) and deep learning (DL) tools, to predict the detection, prognosis, or risk of lung cancer from various medical imaging modalities, including CT, magnetic resonance imaging (MRI), positron emission tomography (PET), and X-rays. The full keyword query used in the search for titles and abstracts is provided in Supplementary eMethods 1a.

For the first search, an initial title and abstract screening was conducted using the **AIScreenR** [34] package in R version 4.4.2 [35], which queried the GPT-4o-mini model [36] ten times independently (eMethods 1b). Studies were included for full-text screening if selected in at least four of the ten queries, which is the default **AIScreenR** probability threshold. The full-text screening was then conducted by screener 1. In the second supplementary search, both title/abstract and full-text screenings were conducted manually by screener 2.

### Data extraction and outcome endpoints

For each study, screeners 1 and 2 extracted information on bibliographic details (authors and year of publication, study setting and geography), study design, data sources, model design and architecture, evaluation metrics, practices for uncertainty quantification (none; evaluation metric-level; model/parameter-level; prediction-level), data/code availability, and use of reporting frameworks for developing clinical prediction models. We reported sample sizes for the training set, the internal hold-out test set, and any external test sets. In the absence of an internal hold-out test set, we recorded k-fold cross-validation details as part of the internal validation. However, if a study employed both cross-validation and an internal hold-out test set, we restricted our data extraction to the hold-out test set. We prioritized extracting the number of participants in the hold-out test sets because they more closely approximate model performance in a fixed test population.

We classified outcome endpoints into three categories: *detection*, which evaluates whether a patient has lung cancer based on the presence or absence of detectable nodules in scans; *prognosis*, which assesses outcomes such as survival or likelihood of recurrence in patients diagnosed with lung cancer; and *risk prediction*, which estimates the risk of developing lung cancer among individuals who have negative images at baseline. Studies with prognosis and risk prediction as endpoints are longitudinal in design, assessing future cancer recurrence or incidence over a defined follow-up period.

### Quality assessment of included studies

Within the same set of included studies, we conducted an embedded, exploratory analysis comparing quality assessments from human reviewers, GenAI tools, and a guideline-based checklist.

### Human scoring

Two statisticians and two clinicians independently rated each study on a scale of 1 to 10, with 10 indicating the highest quality. There were no prescribed guidelines as this was intended as an exploratory overall impression measure. Thus, reviewers set their self-defined criteria before starting scoring, and these criteria are reported in eMethods 2. Statisticians focused on clarity and rigor of reporting, methodological design, and overall reproducibility, while clinicians focused on clinical clarity, follow-up time, outcomes, model performance, and applicability to the patient groups of interest.

### AI-based scoring

Two AI agents (GPT-5 and Gemini 2.5 Pro) were prompted to generate and apply evaluation criteria emphasizing scientific rigor, validation, clinical relevance, and generalizability of published research papers from January 1, 2023, to June 30, 2025, on prediction models proposed for lung cancer detection, prognosis, and prediction from imaging data. Each AI agent scored all included studies on a 1 to 10 scale in five independent runs, and the averaged scores were derived and used for comparison and correlation analysis. We developed the prompts collaboratively among the study team, and the details of the prompting procedure are in eMethods 3. To assess whether role/persona assignment and assuming a specific expert profile changed scoring, each AI agent was instructed to adopt two personas: (a) the perspective of an M.D.-level and board-certified radiologist who screens for lung cancer in the United States or (b) a Ph.D.-level statistician with 10 years of working experience in the United States (eMethods 4).

### Established guideline-based scoring

Each included study was also critically appraised by screener 2 using items from the CHARMS guideline [28]. Scores were standardized to a 10-point scale, where higher scores indicate lower risk of bias. Non-applicable items in the checklist included “outcome assessed without knowledge of candidate predictors,” “candidate predictors part of the outcome,” and “predictors assessed blinded for outcome or each other.” The itemized checklist is provided in Supplementary eTable 1.

### Correlation analysis

To compare the agreement of quality assessment scores across statisticians, clinicians, AI agents, and the CHARMS checklist, we computed pairwise Spearman rank correlations and Pearson correlations to evaluate both rank and linear concordance.

## Results

### Search results

The initial OpenAlex query yielded 838 publications. Of these, we excluded 319 publications because their abstracts were not accessible through OpenAlex. Using the **AIScreenR** package, the GPT-4o-mini model identified 77 candidate studies for full-text screening. From the first search period (January 1, 2023-March 27, 2025), 35 studies were included after full-text screening. The second search, covering the period from March 27 to June 30, 2025, yielded 1 additional study, bringing the total to 36 studies for qualitative synthesis. A detailed flow diagram of the searches is shown in Figure 1.

**Figure 1.**
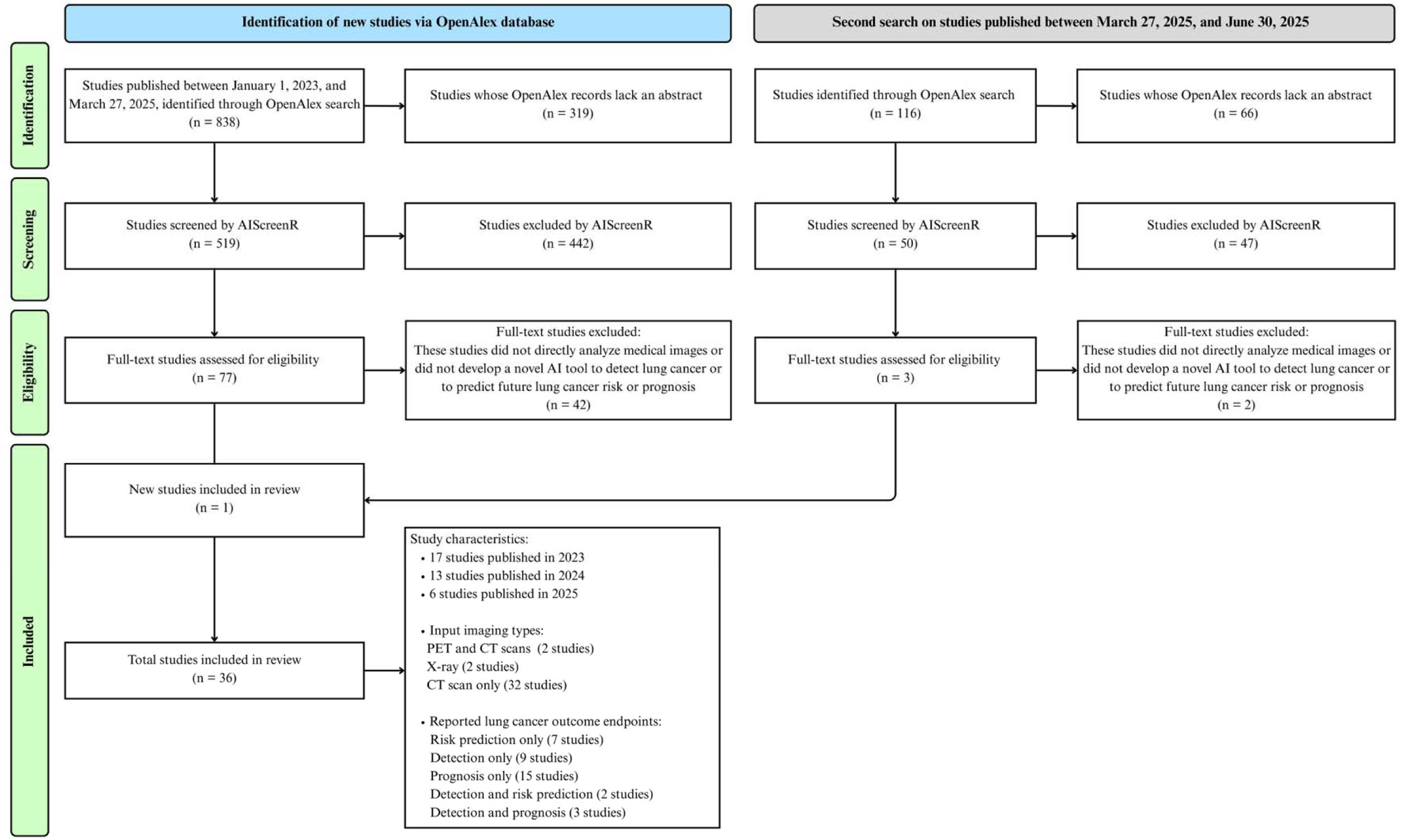
Flow diagram of first search from January 1, 2023, to March 27, 2025 (inclusive), and the second search from March 27 to June 30, 2025 (inclusive), for the cross-sectional meta-research study

### Study characteristics and predicted outcome endpoints

Among the 36 included studies, 17 studies (47%) were published in 2023, 13 studies (36%) in 2024, and 6 studies (17%) in 2025. The input modalities included only CT scans in 32 studies, X-rays in 2 studies, and both PET and CT scans in 2 studies. Reported lung cancer outcome endpoints consisted of only detection in 9 studies, only prognosis in 15 studies, only risk prediction in 7 studies, both detection and risk prediction in 2 studies, and both detection and prognosis in 3 studies. Table 1 provides a detailed summary of study-level characteristics, including first author and publication date, algorithm type, input image modality, whether additional input data were used, outcome endpoints, data source, test set source, reported uncertainty quantification, data/code availability, average statistician and clinician quality scores, average AI agent quality scores (GPT-5, Gemini 2.5 Pro; averaged over five runs), and the CHARMS-based score (all scaled out of 10).

**Table 1.** Summary of study characteristics from the 36 included studies. **Abbreviations:** Demographics (Demo.), Computed Tomography (CT), Low-Dose Computed Tomography (LDCT), Positron Emission Tomography (PET), Reported uncertainty quantification of either evaluation metrics, effect estimates hazard ratios or odds ratios, or error-free survival probabilities (Reported UQ). **Algorithm type abbreviations:** Convolutional Neural Network (CNN), Conditional Generative Adversarial Network (cGAN), Decision Tree (DT), Gradient Boosting Machine (GBM), Generalized Linear Model (GLM), Logistic Regression (LR), Random Forest (RF), Recurrent Neural Network (RNN), Support Vector Machine (SVM), Vision Transformer (ViT), Extreme Gradient Boosting (XGBoost).

| Authors<br>Publication<br>Date | Algorithm<br>Type | Input Image | Input Data |  | Outcome<br>Endpoints | Data<br>Source | Test Set Source |  | Reported UQ | Availability of |  | Human subjective assessment<br>average scores (out of 10) |  | AI agents average scores<br>(out of 10) |  | CHARMS quality<br>score (out of 10) |
| --- | --- | --- | --- | --- | --- | --- | --- | --- | --- | --- | --- | --- | --- | --- | --- | --- |
|  |  |  | Demo. | Clinical |  |  | Internal | External |  | Data | Code | Statisticians | Clinicians | GPT-5 | Gemini 2.5<br>Pro |  |
| Aslani<br>et al. [37]<br>May 2024 | CNN, RNN | LDCT scan,<br>clinical data | ✓ | ✓ | Risk<br>prediction | North<br>America | X | ✓ | None | ✓ | X | 7.40 | 7.0 | 7.22 | 8.2 | 8.3 |
| Christie et al.<br>[38]<br>May 2025 | CNN | CT and PET<br>scans | ✓ | ✓ | Prognosis | North<br>America | ✓ | ✓ | Hazard<br>ratios | X | X | 7.65 | 7.0 | 7.70 | 8.8 | 9.3 |
| Ebrahimpour<br>et al. [39]<br>October 2024 | GBM and<br>XGBoost,<br>SVM, RF, LR | Radiomic<br>features from<br>CT scans | X | X | Risk<br>prediction | North<br>America | ✓ | X | Evaluation<br>metrics | ✓ | X | 6.40 | 8.0 | 6.10 | 3.8 | 9.1 |
| Eldho<br>et al. [40]<br>February 2024 | CNN | CT scans | X | X | Detection | Asia | ✓ | X | None | X | X | 3.75 | 5.5 | 2.96 | 1.0 | 6.9 |
| Fanizzi<br>et al. [41]<br>November<br>2023 | ViT | CT scan<br>slice | X | X | Prognosis | North<br>America | X | X | Evaluation<br>metrics | ✓ | ✓ | 6.50 | 7.0 | 5.60 | 5.2 | 8.6 |
| Gainey<br>et al. [42]<br>April 2023 | CNN, LR | CT scans | X | X | Prognosis | North<br>America | X | ✓ | Hazard<br>ratios | X | X | 6.90 | 8.5 | 6.42 | 7.4 | 8.3 |
| Hao et al. [43]<br>November<br>2024 | CNN,<br>XGBoost | CT scan<br>regions of<br>interest,<br>clinical data | ✓ | ✓ | Prognosis | Asia | ✓ | X | None | X | X | 6.90 | 8.5 | 6.14 | 5.8 | 8.6 |
| Hermoza et al.<br>[44]<br>July 2024 | CNN | Chest X-ray | X | X | Detection,<br>prognosis | North<br>America | ✓ | X | None | ✓ | X | 6.90 | 8.0 | 5.86 | 6.2 | 6.6 |

| Authors<br>Publication Date | Algorithm<br>Type | Input Image | Input Data |  | Outcome<br>Endpoints | Data Source | Test Set Source |  | Reported<br>UQ | Availability of |  | Human subjective assessment<br>average scores (out of 10) |  | AI agents average scores<br>(out of 10) |  | CHARMS<br>quality score<br>(out of 10) |
| --- | --- | --- | --- | --- | --- | --- | --- | --- | --- | --- | --- | --- | --- | --- | --- | --- |
|  |  |  | Demo. | Clinical |  |  | Internal | External |  | Data | Code |  |  |  |  |  |
|  |  |  |  |  |  |  |  |  |  |  |  | Statisticians | Clinicians | GPT-5 | Gemini 2.5<br>Pro |  |
| **Hu et al. [45]**<br>September 2023 | CNN | PET and CT scans, and combined images | X | X | Prognosis | Asia | √ | X | Evaluation metrics | √ | X | 4.90 | 7.5 | 5.58 | 3.2 | 7.9 |
| **Huang et al. [46]**<br>March 2024 | Ensemble (SVM, LR, RF) | Radiomic features from CT scans, clinical data | X | √ | Prognosis | Asia | X | √ | Odds ratios | X | X | 8.00 | 7.5 | 7.80 | 8.4 | 9.0 |
| **Kim et al. [47]**<br>May 2023 | CNN | 2D slices of CT scans | X | X | Prognosis | North America, Asia | X | X | Evaluation metrics | √ | √ | 7.75 | 7.5 | 6.14 | 5.6 | 7.9 |
| **Liu et al. [48]**<br>June 2024 | CNN | CT scan image patches | X | X | Prognosis | Asia | √ | √ | Evaluation metrics | X | X | 6.65 | 8.0 | 6.82 | 8.6 | 9.0 |
| **Mahajan et al. [49]**<br>February 2025 | CNN | CT scans | X | X | Detection | North America, Asia | √ | X | Evaluation metrics | √ | X | 6.50 | 7.5 | 6.02 | 7.2 | 6.6 |
| **Maijeddah et al. [50]**<br>December 2024 | SVM | CT scans | X | X | Detection | Asia | √ | X | None | X | X | 5.25 | 6.5 | 3.34 | 1.4 | 8.6 |
| **Mikhael et al. [16]**<br>January 2023 | CNN | LDCT scans | X | X | Risk prediction | North America, Asia | √ | √ | Evaluation metrics | √ | √ | 9.75 | 8.5 | 8.74 | 10.0 | 10.0 |
| **Mu et al. [51]**<br>May 2023 | CNN | CT scans | X | X | Detection | Asia | √ | X | None | X | X | 5.65 | 8.0 | 6.20 | 6.8 | 8.3 |
| **Ottaiano et al. [52]**<br>June 2024 | SVM | Radiomic features from CT scans | X | X | Prognosis | Europe | X | X | Hazard ratios | X | X | 4.90 | 7.0 | 4.76 | 3.2 | 8.6 |
| **Paetz et al. [53]**<br>April 2023 | CNN | Longitudinal LDCT scans | X | X | Risk prediction | North America | X | X | Error-free survival probabilities | X | X | 5.65 | 7.0 | 5.84 | 4.8 | 8.6 |
| **Park et al. [54]**<br>July 2024 | CNN | CT scans | X | X | Prognosis | North America | √ | X | None | X | X | 6.00 | 7.5 | 3.84 | 2.4 | 7.9 |
|  |  |  | Demo. | Clinical |  |  | Internal | External |  | Data | Code |  |  |  |  |  |
|  |  |  |  |  |  |  |  |  |  |  |  | Statisticians | Clinicians | GPT-5 | Gemini 2.5<br>Pro |  |
| **Prabakaran et al. [55]**<br>February 2023 | CNN | CT scans | X | X | Detection, prognosis | North America | √ | X | None | √ | X | 5.25 | 6.5 | 3.40 | 1.8 | 7.2 |
| **Raza et al. [56]**<br>September 2023 | CNN | CT scans | X | X | Detection | Asia | √ | X | None | √ | X | 4.75 | 7.0 | 3.86 | 1.8 | 6.9 |
| **Salehjahromi et al. [57]**<br>March 2024 | cGAN, XGBoost, RF, SVM, autoencoder | CT scans | X | X | Detection, Risk prediction | North America | √ | √ | Evaluation metrics | √ | √ | 9.15 | 9.0 | 7.62 | 10.0 | 8.6 |
| **Sousa et al. [58]**<br>June 2023 | CNN, RF | CT scan regions of interest, clinical data | √ | √ | Detection | North America | √ | X | Evaluation metrics | √ | X | 5.75 | 7.0 | 5.42 | 5.2 | 8.3 |
| **Tang et al. [59]**<br>July 2023 | Ensemble (SVM, RF, DT, XGBoost, GLM) | Radiomic features from CT scans, clinical data | √ | √ | Prognosis | North America | √ | X | Evaluation metrics | √ | X | 5.75 | 7.0 | 5.70 | 5.6 | 7.8 |
| **Tonneau et al. [60]**<br>July 2023 | CNN | CT scans, clinical data | √ | √ | Prognosis | North America | X | √ | Evaluation metrics | X | X | 5.90 | 7.5 | 7.42 | 9.0 | 8.3 |
| **Vemula et al. [61]**<br>May 2024 | CNN | CT scans | √ | X | Detection | North America | √ | X | None | √ | X | 4.15 | 6.0 | 2.90 | 1.2 | 6.2 |
| **Verma et al. [62]**<br>July 2024 | Autoencoder | CT scans, radio-genomic data, and clinical data | √ | √ | Risk prediction | North America | X | √ | Evaluation metrics | √ | √ | 8.50 | 7.5 | 7.40 | 8.4 | 7.6 |
| **Wan et al. [63]**<br>February 2025 | GBM, LR | CT scans, clinical data | √ | √ | Detection, prognosis | Asia | √ | X | Evaluation metrics | X | X | 6.25 | 7.5 | 6.70 | 6.6 | 8.6 |

| Authors<br>Publication Date | Algorithm<br>Type | Input Image | Input Data |  | Outcome<br>Endpoints | Data Source | Test Set Source |  | Reported<br>UQ | Availability of |  | Human subjective assessment<br>average scores (out of 10) |  | AI agents average<br>scores (out of 10) |  | CHARMS quality<br>score (out of 10) |
| --- | --- | --- | --- | --- | --- | --- | --- | --- | --- | --- | --- | --- | --- | --- | --- | --- |
|  |  |  | Demo. | Clinical |  |  | Internal | External |  | Data | Code | Statisticians | Clinicians | GPT-5 | Gemini 2.5<br>Pro |  |
| **Wang et al. [64]**<br>July 2023 | CNN | CT scans of lung nodules | X | X | Detection | North America | √ | X | None | √ | √ | 6.90 | 7.5 | 4.08 | 5.0 | 6.9 |
| **Weiss et al. [65]**<br>May 2023 | CNN | Chest X-ray | X | X | Risk prediction | North America | √ | √ | Evaluation metrics | √ | √ | 8.90 | 8.0 | 8.20 | 9.2 | 9.7 |
| **Yang et al. [66]**<br>January 2025 | CNN | CT scans | X | X | Risk prediction | Asia | √ | X | Evaluation metrics | X | X | 7.25 | 8.5 | 7.08 | 7.8 | 9.7 |
| **Yin et al. [67]**<br>January 2025 | CNN | Radiomic features from CT scans, clinical data | √ | √ | Prognosis | Asia | √ | √ | Evaluation metrics | X | X | 5.65 | 7.0 | 7.98 | 8.2 | 9.0 |
| **Zhang et al. [68]**<br>March 2025 | DT, RF, SVM, LR, GBM and XGBoost | CT scans, clinical data | √ | √ | Prognosis | Asia | √ | √ | Evaluation metrics | X | X | 6.50 | 7.5 | 7.52 | 8.4 | 9.7 |
| **Zheng et al. [69]**<br>January 2023 | CNN, LR | CT scan patches, clinical data | √ | √ | Prognosis | Europe | √ | √ | Evaluation metrics | X | X | 5.65 | 6.5 | 7.24 | 7.4 | 7.9 |
| **Zhou et al. [70]**<br>July 2023 | CNN | CT scans with location of nodule | X | X | Detection, Risk prediction | Asia | √ | √ | Evaluation metrics | X | √ | 6.50 | 7.5 | 7.50 | 8.4 | 9.0 |
| **Zyla et al. [71]**<br>December 2023 | LR, RF | Radiomic features from LDCT scans, clinical data | X | √ | Detection | North America, Europe | √ | X | Evaluation metrics | X | X | 5.25 | 7.0 | 5.84 | 5.4 | 7.6 |

### Study populations, study design, use of additional sociodemographic, behavioral or clinical input data in model development (in addition to images)

Frequency distributions of study characteristics were compared across the 36 included studies (Figure 2). Most studies used imaging data from populations in North America (21 studies) or Asia (16 studies), while only 3 studies included data from European populations. There were 4 studies that were double counted in the above numbers, where 3 studies had study populations from both North America and Asia, and 1 study had populations from both North America and Europe. The vast majority were retrospective (35/36, 97%, Supplementary eFigure 1). Only one study was prospective, evaluating lung cancer prognosis, which spanned from December 2020 to April 2022 and included all sequentially diagnosed and radiologically staged lung cancer patients treated at the University of Naples Federico II [52].

**Figure 2.**
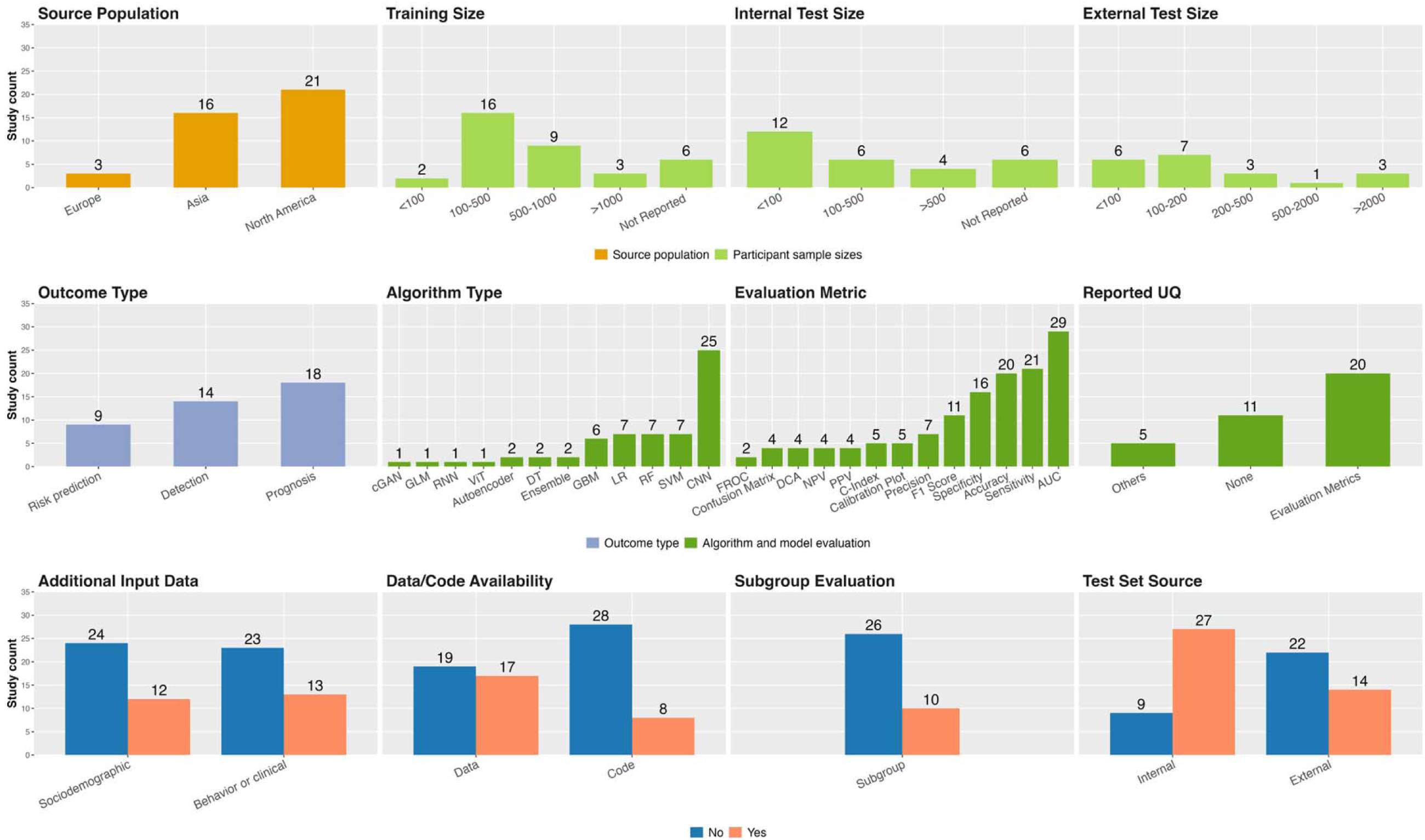
Summary characteristics of the 36 included studies. Abbreviations include UQ for uncertainty quantification. Algorithm type abbreviations are Convolutional Neural Network (CNN), Decision Tree (DT), conditional Generative Adversarial Network (cGAN), Gradient Boosting Machine (GBM), Generalized Linear Model (GLM), Logistic Regression (LR), Random Forest (RF), Recurrent Neural Network (RNN), Support Vector Machine (SVM), Vision Transformer (ViT). Evaluation metric abbreviations are area under the curve (AUC), concordance index (C-index), positive predictive value (PPV), negative predictive value (NPV), decision curve analysis (DCA), free-response receiver operating characteristic (FROC).

Covariates appeared in two separate roles: inputs to the model or stratification factors for subgroup assessment. There were 12 studies (33%) that included sociodemographic variables such as age, sex, or social determinants, and 13 studies (36%) that incorporated behavioral or clinical variables such as smoking status or medical history. Subgroup performance was reported in 10 of 36 studies (e.g., by sex, smoking status, tumor stage, or nodule characteristics). Of these, 4 studies used covariates both as model inputs and as stratification factors for subgroup analysis, whereas 6 studies used the covariates only for subgroup analysis and did not include them as predictors in the model.

### Sample sizes of training, internal test set, and external test set

Training and testing sample sizes of participants varied widely across studies. Across the 36 studies, training set sizes ranged from 57 to 40,643 participants (reported by 30 studies). Of the remaining six studies, five reported only image-level sample sizes (e.g., numbers of scans, not participants), and one did not report any training set size information. Internal test sets ranged from 35 to 10,155 participants (reported by 21 of 27 studies). There were eight studies that only used cross-validation instead of any internal hold-out test. At least one external test set was reported in 14 of the 36 included studies (39%), including four that used multiple external test sets [16,57,62,65]. Among these 14 studies, external test sizes ranged from 26 to 10,567 participants. Considering studies that reported exact counts, the typical train:internal split was roughly 4:1, and the train:external ratio was also about 4:1. Detailed distributions of training, internal, and external test sample sizes of participants are provided in Supplementary eResults 1. *Model types and architectures, pre-trained model specifications, and reporting of model parameters*

The majority of studies, 25 of 36 (69%), utilized CNN to analyze imaging data. Other commonly applied methods included support vector machines (SVM), random forests (RF), and logistic regression (LR), each used in 7 studies, and gradient boosting machines (GBM), used in 6 studies. Less commonly employed methods included ensemble learning, decision trees (DT), and autoencoders (2 studies each), and conditional generative adversarial network (cGAN), generalized linear models (GLM), recurrent neural network (RNN), and vision transformer (ViT), each appearing in a single study. Because several studies evaluated more than one modeling approach, these study counts are not mutually exclusive and sum to more than the total number of studies.

Autoencoders were used for feature extraction rather than direct classification. In Salehjahromi et al. [57], autoencoders were used to extract latent features from PET scans prior to training a random forest classifier. In Verma et al. [62], they were applied to identify prognostic gene biomarkers and molecular pathways for building a Cox proportional hazards model. Several studies combined multiple machine learning methods to make lung cancer predictions, e.g., CNN to classify 3D CT nodule region of interest and RF algorithm to classify clinical data [58], CNN for building an imaging model and LR for building a clinical model [69], and similarly GBM and LR for incorporating imaging and clinical data [63], respectively. Model-type patterns were broadly similar across detection, prognosis, and risk prediction endpoints with no clear clustering of specific algorithms by endpoint (eTable 2).

To characterize model configurations, we extracted information on pre-trained architectures and final model parameters of 25 studies that used CNNs in their model development. Sixteen of the 25 studies used pre-trained models during feature extraction, selection, or prediction, leveraging representations learned from large external datasets (e.g., ImageNet) to improve performance on the study-specific imaging data and reduce training time. Only 6 out of 25 studies reported the number of parameters in their final models. Details of the pre-trained model architectures and final model parameters of these 25 studies are available in eTable 3 and eFigure 2, respectively.

### Handling of class imbalance during model training

Class imbalance, in which the dataset contains substantially more cases without the outcome (e.g., lung cancer negative class) than with the outcome (e.g., lung cancer positive class), was unaddressed in 22 studies (61%), which may have relied on threshold adjustments during validation or testing. In settings where the number of negative cases substantially exceeds the number of positive cases, explicit imbalance-handling strategies during training are generally recommended to avoid models that preferentially learn the majority class and perform poorly for the minority class [72,73]. Fourteen studies (39%) applied imbalance-handling techniques during training, including minority-target augmentation (3 studies) [50,55,56], synthetic minority over-sampling technique or SMOTE (2 studies) [39,62], any adjustments in convolutional neural networks (2 studies) [40,69], and other methods reported in Supplementary eTable 4.

### Performance metrics and uncertainty quantification

The most commonly reported evaluation metric was the area under the curve (AUC), used in 29 studies. Other frequently reported metrics included sensitivity (21 studies), accuracy (20 studies), and specificity (16 studies). Uncertainty quantification was reported in 25 (69%) out of the 36 included studies, encompassing different types of outcomes. Specifically, 20 studies quantified uncertainty in model evaluation metrics, e.g., confidence intervals for AUC, sensitivity, or accuracy; four studies quantified uncertainty in effect estimates, e.g., confidence intervals for hazard ratios or odds ratios; and 1 study reported uncertainty in predicted survival probabilities.

### Subgroup level evaluation

Increasing lung cancer incidence among younger adults, women, and non-smokers is being recognized. Thus, subgroup analysis is important to evaluate whether AI models perform well across evolving risk profiles. Yet, only 10 out of 36 studies (28%) evaluated model performance across population subgroups. Within these 10 studies, some stratified the study population by age, biological sex, race, and smoking status [16,43,65]. As most models were trained on racially homogeneous or non-probability sampled cohorts, these models may underperform in subpopulations, and generalizability to specific sub-populations of interest can still be unreliable.

### Study transparency and reproducibility

Seventeen studies (47%) used datasets described as publicly available, though accessibility varied. Some studies used open-access datasets that required no permission, such as The Cancer Imaging Archive, while others used controlled-access datasets that required formal access requests and approval (e.g., NLST or institutional datasets). Only 8 studies (22%) provided publicly available code.

### Quality assessment comparison between statisticians, clinicians, GPT-5, Gemini 2.5 Pro, and existing guideline-based (CHARMS) scoring

The median quality scores, on a 1-10 scale, were 7 and 5 for statisticians 1 and 2; 8 and 7 for clinicians 1 and 2; 6.14 and 6.4 for GPT-5 and Gemini 2.5 Pro; and 8.3 for CHARMS-based checklist. Pairwise Spearman rank correlations were calculated for quality scores evaluated by statisticians 1 and 2, clinicians 1 and 2, AI agents, and the CHARMS-based checklist (Figure 3). There was notable variation between statisticians and clinicians: correlations between clinician 1 and statisticians 1 and 2 were relatively weak (Spearman’s rank correlation coefficient p = 0.28 and 0.15, respectively) compared with a stronger correlation between clinician 2 and statisticians 1 and 2 (p = 0.81 and 0.70, respectively). Variation was also observed within the two clinicians’ scores, shown by a weak correlation (p = 0.22), whereas statisticians’ evaluations were more correlated with each other (p = 0.57) compared to those of clinicians.

**Figure 3.**
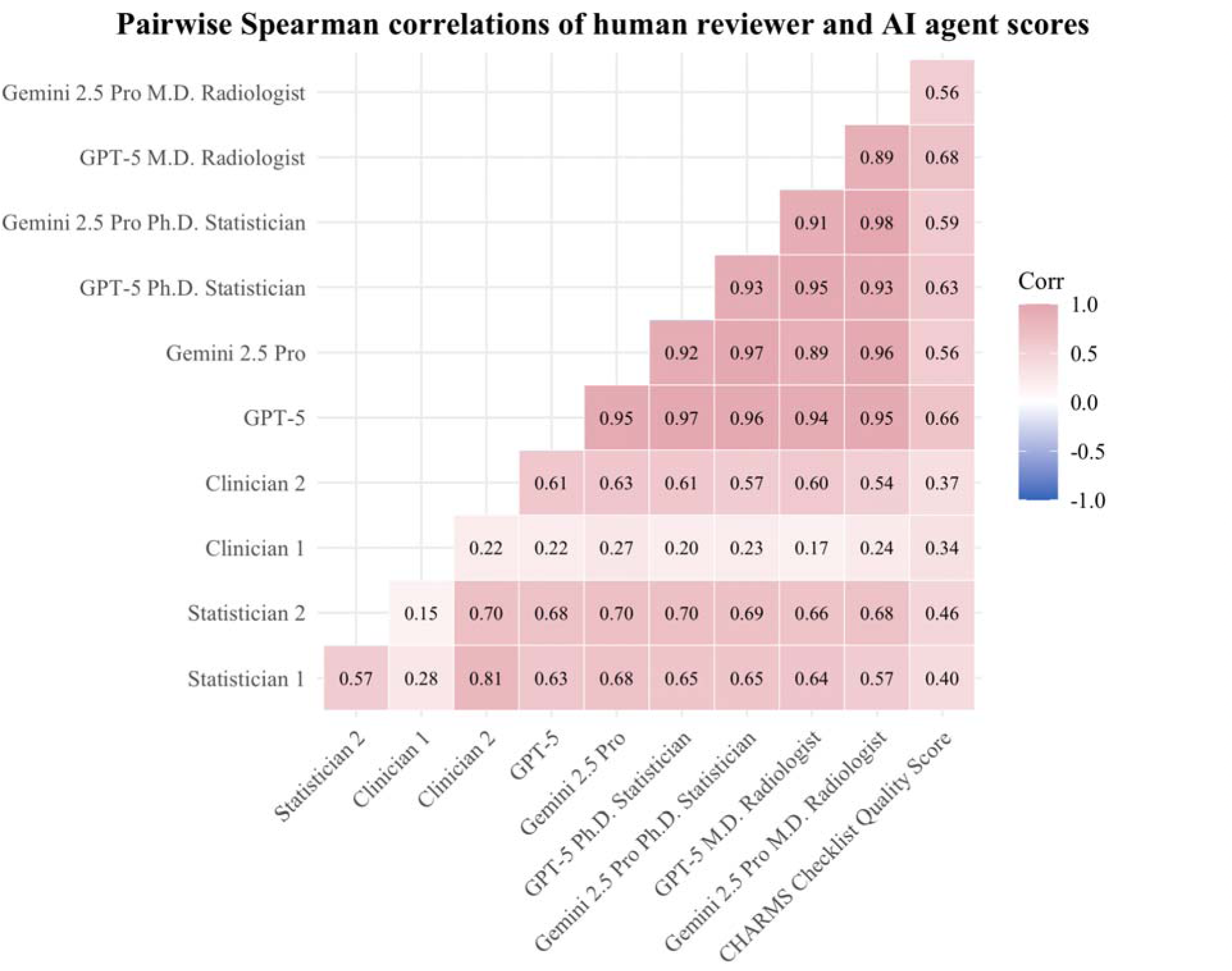
Pairwise Spearman correlation plot on quality scorings, with higher correlations being represented by darker red shades and negative correlations by progressively darker blue shades. The scores include those by statisticians 1 and 2, clinicians 1 and 2, GPT-5 and Gemini 2.5 Pro without assigned roles averaged over five runs, and GPT-5 and Gemini 2.5 Pro assigned as M.D.-level radiologist averaged over five runs, and GPT-5 and Gemini 2.5 Pro assigned as Ph.D.-level statistician averaged over five runs.

AI agents were highly consistent, with correlations ≥ 0.89 across models and role assignments. Statisticians’ scores correlated more strongly with AI agents than did clinicians’ scores, suggesting greater alignment in methodological assessment. Gemini 2.5 Pro had the greatest correlation of study scores with both statisticians 1 and 2 ( p = 0.68 and 0.70, respectively), followed by GPT-5 (p = 0.65 and 0.70, respectively) and Gemini 2.5 Pro with Ph.D. statistician role (p = 0.65 and 0.69, respectively).

Correlations with the CHARMS-based scoring were weaker for statisticians and clinicians (p ≤ 0.46) than for all AI agents, with GPT-5 (no role p = 0.66, Ph.D. statistician p = 0.63, M.D. clinician p = 0.68) showing a stronger correlation compared to Gemini 2.5 Pro (no role p = 0.56, Ph.D. statistician p = 0.59, M.D. clinician p = 0.56). Pairwise Pearson correlations yielded similar patterns (eFigure 3). Correlations between the average scores of two statisticians and two clinicians, scores from the two AI agents, and the CHARMS-based scores are shown in eFigures 4 and 5, and per-study scores for each rater are listed in eTable 5. Examples of the criteria generated and used by GPT-5 and Gemini 2.5 Pro are provided in eResults 2-7. Overlap between evaluation criteria from CHARMS checklist and both GPT-5 and Gemini 2.5 Pro without role assignment is described in eResults 8.

**Figure 4.**
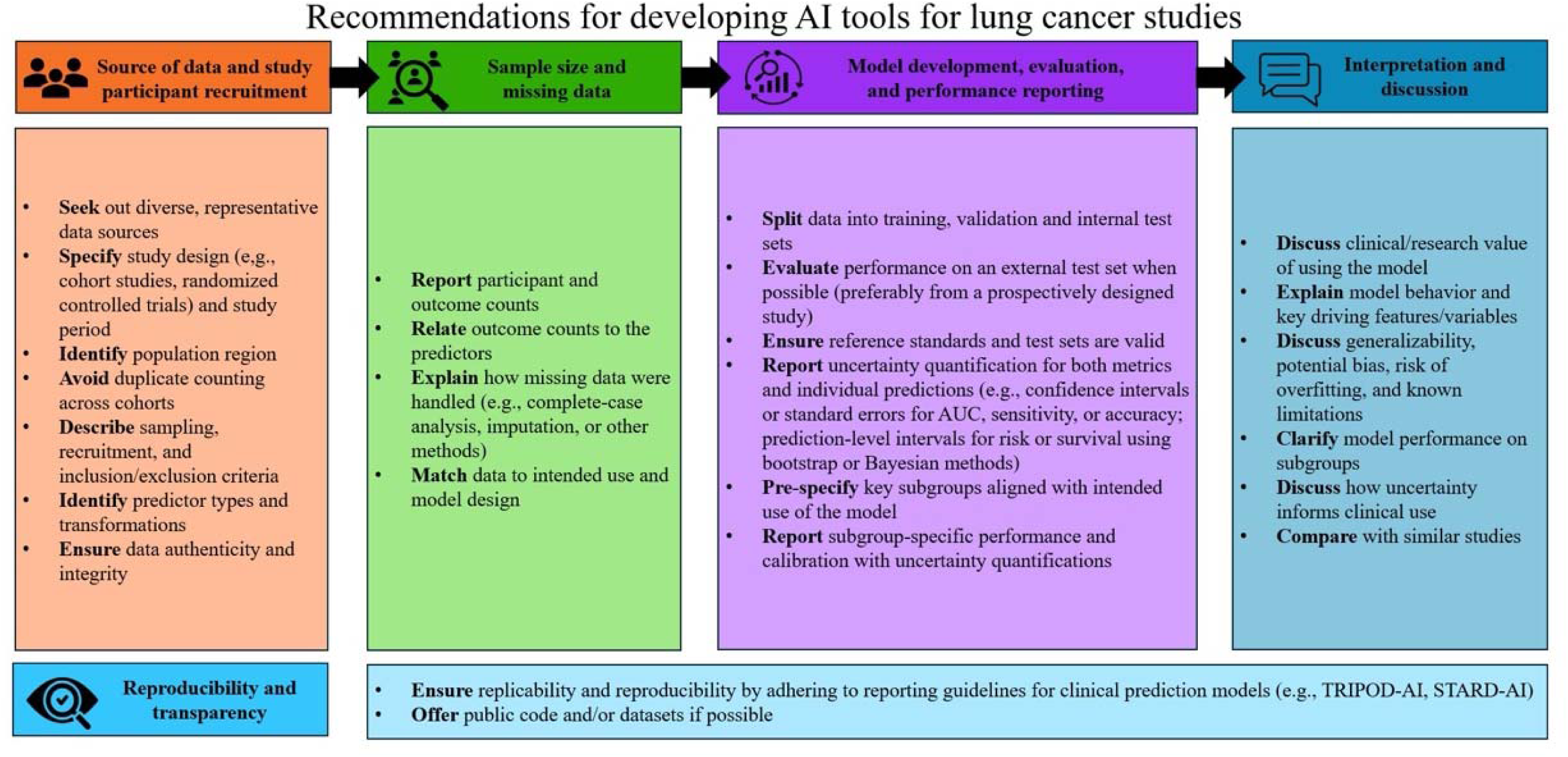
Proposed best practices for developing and evaluating AI tools for lung cancer studies, including key sections of source of data and study participant recruitment, sample size and missing data, model development, evaluation, and performance reporting, interpretation and discussion, and reproducibility and transparency.

## Discussion

### Current landscape of recent studies developing and using AI-based prediction models for lung cancer outcomes

This meta-research study describes and critically evaluates current practices in 36 recent studies developing AI-based prediction models for lung cancer detection, prognosis, and risk prediction. A key contribution of this meta-research study is the identification of a transparency gap in the architectural and statistical design of current lung cancer prediction models. While the field is eager to adopt AI models, our analysis of algorithm types reveals a stagnation in architectural diversity. The vast majority (69%) of studies relied on CNNs, whereas only a small number of studies explored alternative architectures such as ViT, cGANs, or autoencoders. Our goal was to document which architectures are currently being used in practice, rather than to assume that these newer approaches would necessarily perform better than CNN-based models. More critically, we observed a widespread lack of reporting on the structural design parameters among the 25 studies that used CNNs, which amplifies the concerns about model opacity and interpretability.

Another major finding is the disconnect between model evaluation and clinical utility regarding uncertainty quantification. AUC serves as a popular metric because it provides a single, threshold-independent score evaluating predictive models, but it does not convey information about calibration or the clinical decision thresholds at which a model would be used. While 69% of studies reported some form of uncertainty quantification, the majority were limited to reporting confidence intervals for global performance metrics (e.g., AUC, sensitivity, and accuracy). Studies focusing on prognosis or risk prediction should quantify and report prediction/risk-level uncertainty, as clinicians need to know not only the model’s accuracy and its uncertainty but also the prediction interval for a specific patient’s risk score or survival probability. As noted in our results, only one out of the 36 included studies provided a similar level of granularity.

Class imbalance handling during training and participant sample sizes of training, internal, and external test sets were highly heterogeneous. The limited use of subgroup reporting (only 28%) suggests a risk of calibration drift and reduced transportability to the population of interest. Furthermore, the lack of prospective studies or randomized clinical trials (RCTs) raises concerns about how in-depth these AI models were tested. As Elemento et al. [74] emphasized, the use of prospective RCTs is crucial for validating the clinical utility and safety of these AI models prior to widespread adoption in cancer care. Limited code sharing and mixed data access further impede verification and reuse of these AI models.

Based on these findings, we summarize key considerations that researchers should incorporate into their workflow when developing AI tools for lung cancer studies in Figure 4, which include assessing data source and participant recruitment, sample size and missing data, model evaluation and performance reporting, interpretation and discussion, and ensuring reproducibility and transparency throughout this workflow.

### Adherence to reporting guidelines among recent predictive studies and AI agent evaluation of study quality

Although several guidelines exist for prediction model development and reporting, adoption of these guidelines was rare. Among the 36 included studies, only one explicitly referenced the TRIPOD statement [75] during model development. In April 2024, an updated statement by TRIPOD-AI aims to promote the complete, accurate, and transparent reporting of prediction model studies regarding the rise of AI methods in prediction models [76]. To support the rapidly expanding field of oncology and AI research, Elemento et al. [74] outlined key actions for integrating AI effectively into oncology research. Ong et al. [77] proposed a bioethical framework for the responsible use of GenAI, particularly large language models, in medicine.

Within this context, our embedded AI-based quality appraisal provides an illustration of how large language models might complement human assessment. Comparing quality scoring by statisticians, clinicians, AI agents (GPT-5 and Gemini 2.5 Pro), and an established guideline-based tool (CHARMS) highlighted both the subjectivity of human evaluations and the potential of automated tools to provide more structural, checklist-driven appraisals. These findings suggest that large language models could support researchers by offering consistent, guideline-based evaluations that streamline study appraisal by humans in literature reviews and encourage better reporting practices and alignment with reporting guidelines for future AI-based lung cancer research. As the research community shifts towards AI agents as co-researchers and co-reviewers, exemplified by the recent Agents4Science conference findings [78], we demonstrate how to approach using AI as a useful tool in developing prediction models for lung cancer outcomes.

## Conclusion

This cross-sectional meta-research study provides a comprehensive synthesis of recent studies that have developed AI tools for predicting lung cancer outcomes. Our findings highlight the need for future studies to use guidelines for prediction model development and improve model evaluation via external test sets and subgroup analyses. The contrast between the technical advances of emerging AI models and the inconsistency in model development and reporting practices underscores the continuing need for standardized and transparent research frameworks in this field.

## Supporting information

Supplementary_eTable5

Conceptualization: BM

Methodology: BM, TW, YH, CY

Supervision: BM, DB

Paper screeners: TW, YH

Statistician and clinician reviewers: TW, CY, GC, EJ Writing – original draft: YH, CY, TW

Writing – review & editing: BM, YH, TW, CY, EJ, GC, DB

## Data Availability

All publication data are publicly available and can be retrieved online from the OpenAlex web application.

## Acknowledgements

We would like to acknowledge Kate Nyhan, MLS IPI PMC, the Research and Education Librarian for Public Health, Cushing/Whitney Medical Library, Yale University, for her feedback and support to the paper.

## References

[1] OpenAI. Introducing ChatGPT. OpenAI. November 30, 2022. Accessed August 15, 2025. https://openai.com/blog/chatgpt.

[2] Sengar SS, Hasan AB, Kumar S, Carroll F. Generative artificial intelligence: a systematic review and applications. Multimedia Tools Appl. 2025;84:23661–23700. doi:10.1007/s11042-024-18473-6.

[3] Zhu E, Muneer A, Zhang J, et al. Progress and challenges of artificial intelligence in lung cancer clinical translation. NPJ Precis Oncol. 2025;9(1):210. Published 2025 Jul 1. doi:10.1038/s41698-025-00986-7.

[4] Jiang X, Hu Z, Wang S, Zhang Y. Deep Learning for Medical Image-Based Cancer Diagnosis. Cancers (Basel). 2023;15(14):3608. Published 2023 Jul 13. doi:10.3390/cancers15143608.

[5] Søgaard A. On the Opacity of Deep Neural Networks. Canadian Journal of Philosophy. 2023;53(3):224–239. doi:10.1017/can.2024.1.

[6] Alzubaidi L, Zhang J, Humaidi AJ, et al. Review of deep learning: concepts, CNN architectures, challenges, applications, future directions. J Big Data. 2021;8(1):53. doi:10.1186/s40537-021-00444-8.

[7] Khan A, Sohail A, Zahoora U, Qureshi AS. A survey of the recent architectures of deep convolutional neural networks. Artificial Intelligence Review. 2020;53:5455–5516. doi:10.1007/s10462-020-09825-6.

[8] Powell S. Lung cancer kills more people worldwide than other cancer types. American Cancer Society. Published July 29, 2024. Accessed August 21, 2025. https://www.cancer.org/research/acs-research-news/lung-cancer-kills-more-people-worldwide-than-other-cancer-types.html.

[9] Kanan M, Alharbi H, Alotaibi N, et al. AI-Driven Models for Diagnosing and Predicting Outcomes in Lung Cancer: A Systematic Review and Meta-Analysis. Cancers (Basel). 2024;16(3):674. Published 2024 Feb 5. doi:10.3390/cancers16030674.

[10] Hammad M, ElAffendi M, El-Latif AAA, Ateya AA, Ali G, Plawiak P. Explainable AI for lung cancer detection via a custom CNN on CT images. Sci Rep. 2025;15(1):12707. Published 2025 Apr 13. doi:10.1038/s41598-025-97645-5.

[11] Dodia S, Annappa B, Mahesh PA. Recent advancements in deep learning based lung cancer detection: A systematic review. Eng Appl Artif Intell. 2022;116:105490. doi:10.1016/j.engappai.2022.105490.

[12] Shao J, Feng J, Li J, Liang S, Li W, Wang C. Novel tools for early diagnosis and precision treatment based on artificial intelligence. Chin Med J Pulm Crit Care Med. 2023;1(3):148–160. doi:10.1016/j.pccm.2023.05.001.

[13] Mehri-Kakavand G, Mdletshe S, Wang A. A Comprehensive Review on the Application of Artificial Intelligence for Predicting Postsurgical Recurrence Risk in Early-Stage Non-Small Cell Lung Cancer Using Computed Tomography, Positron Emission Tomography, and Clinical Data. J Med Radiat Sci. Published online January 23, 2025. doi:10.1002/jmrs.860.

[14] Lococo F, Ghaly G, Chiappetta M, et al. Implementation of Artificial Intelligence in Personalized Prognostic Assessment of Lung Cancer: A Narrative Review. Cancers (Basel). 2024;16(10):1832. Published 2024 May 10. doi:10.3390/cancers16101832.

[15] Raghu VK, Walia AS, Zinzuwadia AN, et al. Validation of a Deep Learning-Based Model to Predict Lung Cancer Risk Using Chest Radiographs and Electronic Medical Record Data. JAMA Netw Open. 2022;5(12):e2248793. Published 2022 Dec 1. doi:10.1001/jamanetworkopen.2022.48793.

[16] Mikhael PG, Wohlwend J, Yala A, et al. Sybil: A Validated Deep Learning Model to Predict Future Lung Cancer Risk From a Single Low-Dose Chest Computed Tomography. J Clin Oncol. 2023;41(12):2191–2200. doi:10.1200/JCO.22.01345.

[17] Chen M, Copley SJ, Viola P, Lu H, Aboagye EO. Radiomics and artificial intelligence for precision medicine in lung cancer treatment. Semin Cancer Biol. 2023;93:97–113. doi:10.1016/j.semcancer.2023.05.004.

[18] National Lung Screening Trial Research Team, Aberle DR, Berg CD, et al. The National Lung Screening Trial: overview and study design. Radiology. 2011;258(1):243–253. doi:10.1148/radiol.10091808.

[19] Li H, Salehjahromi M, Godoy MCB, et al. Lung Cancer Risk Prediction in Patients with Persistent Pulmonary Nodules Using the Brock Model and Sybil Model. Cancers (Basel). 2025;17(9):1499. Published 2025 Apr 29. doi:10.3390/cancers17091499.

[20] Binuya MAE, Engelhardt EG, Schats W, Schmidt MK, Steyerberg EW. Methodological guidance for the evaluation and updating of clinical prediction models: a systematic review. BMC Med Res Methodol. 2022;22(1):316. Published 2022 Dec 12. doi:10.1186/s12874-022-01801-8.

[21] de Hond AAH, Leeuwenberg AM, Hooft L, et al. Guidelines and quality criteria for artificial intelligence-based prediction models in healthcare: a scoping review. NPJ Digit Med. 2022;5(1):2. Published 2022 Jan 10. doi:10.1038/s41746-021-00549-7.

[22] Dhiman P, Ma J, Kirtley S, et al. Prediction model protocols indicate better adherence to recommended guidelines for study conduct and reporting. J Clin Epidemiol. 2024;169:111287. doi:10.1016/j.jclinepi.2024.111287.

[23] Efthimiou O, Seo M, Chalkou K, Debray T, Egger M, Salanti G. Developing clinical prediction models: a step-by-step guide. BMJ. 2024;386:e078276. Published 2024 Sep 3. doi:10.1136/bmj-2023-078276.

[24] Kwong JCC, Khondker A, Lajkosz K, et al. APPRAISE-AI Tool for Quantitative Evaluation of AI Studies for Clinical Decision Support. JAMA Netw Open. 2023;6(9):e2335377. Published 2023 Sep 5. doi:10.1001/jamanetworkopen.2023.35377.

[25] Luo W, Phung D, Tran T, et al. Guidelines for Developing and Reporting Machine Learning Predictive Models in Biomedical Research: A Multidisciplinary View. J Med Internet Res. 2016;18(12):e323. Published 2016 Dec 16. doi:10.2196/jmir.5870.

[26] Tejani AS, Klontzas ME, Gatti AA, Mongan JT, Moy L, Park SH, Kahn CE Jr. Checklist for Artificial Intelligence in Medical Imaging (CLAIM): 2024 Update. Radiol Artif Intell. 2024;6(4):e240300. doi:10.1148/ryai.240300.

[27] U.S. Food and Drug Administration; Health Canada; Medicines and Healthcare products Regulatory Agency. Good Machine Learning Practice for Medical Device Development: Guiding Principles. October 2021. Accessed September 3, 2025. https://www.fda.gov/medical-devices/software-medical-device-samd/good-machine-learning-practice-medical-device-development-guiding-principles.

[28] Moons KG, de Groot JA, Bouwmeester W, et al. Critical appraisal and data extraction for systematic reviews of prediction modelling studies: the CHARMS checklist. PLoS Med. 2014;11(10):e1001744. Published 2014 Oct 14. doi:10.1371/journal.pmed.1001744.

[29] Wolff RF, Moons KGM, Riley RD, et al. PROBAST: A Tool to Assess the Risk of Bias and Applicability of Prediction Model Studies. Ann Intern Med. 2019;170(1):51–58. doi:10.7326/M18-1376.

[30] Sounderajah V, Guni A, Liu X, et al. The STARD-AI reporting guideline for diagnostic accuracy studies using artificial intelligence. Nat Med. 2025. doi:10.1038/s41591-025-03953-8.

[31] OpenAI. ChatGPT (GPT-5) [large language model]. OpenAI; 2025. Accessed September 28, 2025. https://platform.openai.com/.

[32] Google Gemini. Gemini 2.5 Pro [large language model]. Google; 2025. Accessed September 28, 2025. https://gemini.google.com

[33] Priem J, Piwowar H, Orr R. OpenAlex: A fully-open index of scholarly works, authors, venues, institutions, and concepts. ArXiv. https://arxiv.org/abs/2205.01833.

[34] Vembye M. AIScreenR: AI Screening Tools in R for Systematic Reviewing. doi:10.32614/CRAN.package.AIscreenR. 10.32614/CRAN.package.AIscreenR.

[35] R Core Team (2025). R: A Language and Environment for Statistical Computing. R Foundation for Statistical Computing, Vienna, Austria. https://www.R-project.org/.

[36] OpenAI. ChatGPT (GPT-4o-mini) [large language model]. OpenAI; 2025. Accessed July 10, 2025. https://platform.openai.com/.

[37] Aslani S, Alluri P, Gudmundsson E, et al. Enhancing cancer prediction in challenging screen-detected incident lung nodules using time-series deep learning. Comput Med Imaging Graph. 2024;116:102399. doi:10.1016/j.compmedimag.2024.102399.

[38] Christie JR, Romine P, Eddy K, et al. Thorax-encompassing multi-modality PET/CT deep learning model for resected lung cancer prognostication: A retrospective, multicenter study. Med Phys. 2025;52(6):4390–4402. doi:10.1002/mp.17862.

[39] Ebrahimpour L, Després P, Manem VSK. Differential Radiomics-Based Signature Predicts Lung Cancer Risk Accounting for Imaging Parameters in NLST Cohort. Cancer Med. 2024;13(20):e70359. doi:10.1002/cam4.70359.

[40] Eldho KJ, Nithyanandh S. (2024) Lung Cancer Detection and Severity Analysis with a 3D Deep Learning CNN Model Using CT-DICOM Clinical Dataset. Indian Journal of Science and Technology. 17(10): 899–910. 10.17485/IJST/v17i10.3085.

[41] Fanizzi A, Fadda F, Comes MC, et al. Comparison between vision transformers and convolutional neural networks to predict non-small lung cancer recurrence. Sci Rep. 2023;13(1):20605. Published 2023 Nov 23. doi:10.1038/s41598-023-48004-9.

[42] Gainey JC, He Y, Zhu R, et al. Predictive power of deep-learning segmentation based prognostication model in non-small cell lung cancer. Front Oncol. 2023;13:868471. Published 2023 Apr 4. doi:10.3389/fonc.2023.868471.

[43] Hao P, Yu Y, Huang CT, et al. Advancing EGFR mutation subtypes prediction in NSCLC by combining 3D pretrained ConvNeXt, radiomics, and clinical features. Front Oncol. 2024;14:1464555. Published 2024 Nov 15. doi:10.3389/fonc.2024.1464555.

[44] Hermoza R, Nascimento JC, Carneiro G. Weakly-supervised preclinical tumor localization associated with survival prediction from lung cancer screening Chest X-ray images. Comput Med Imaging Graph. 2024;115:102395. doi:10.1016/j.compmedimag.2024.102395.

[45] Hu D, Li X, Lin C, Wu Y, Jiang H. Deep Learning to Predict the Cell Proliferation and Prognosis of Non-Small Cell Lung Cancer Based on FDG-PET/CT Images. Diagnostics (Basel). 2023;13(19):3107. Published 2023 Sep 30. doi:10.3390/diagnostics13193107.

[46] Huang D, Lin C, Jiang Y, et al. Radiomics model based on intratumoral and peritumoral features for predicting major pathological response in non-small cell lung cancer receiving neoadjuvant immunochemotherapy. Front Oncol. 2024;14:1348678. Published 2024 Mar 20. doi:10.3389/fonc.2024.1348678.

[47] Kim G, Park YM, Yoon HJ, Choi JH. A multi-kernel and multi-scale learning based deep ensemble model for predicting recurrence of non-small cell lung cancer. PeerJ Comput Sci. 2023;9:e1311. Published 2023 May 2. doi:10.7717/peerj-cs.1311.

[48] Liu Y, Huang J, Chen JC, Chen W, Pan Y, Qiu J. Predicting treatment response in multicenter non-small cell lung cancer patients based on federated learning. BMC Cancer. 2024;24(1):688. Published 2024 Jun 5. doi:10.1186/s12885-024-12456-7.

[49] Mahajan A, Agarwal R, Agarwal U, et al. A Novel Deep Learning-Based (3D U-Net Model) Automated Pulmonary Nodule Detection Tool for CT Imaging. Curr Oncol. 2025;32(2):95. Published 2025 Feb 8. doi:10.3390/curroncol32020095.

[50] Maijeddah UI, Yusuf SA, Abdullhai M, Hassan IH. A hybrid transfer learning model with optimized SVM using honey badger optimization algorithm for multi-class lung cancer classification. Science World Journal. 2025;19(4):977–986. Published 2025 February 14. doi: 10.4314/swj.v19i4.10.

[51] Mu J, Kuang K, Ao M, et al. Deep learning predicts malignancy and metastasis of solid pulmonary nodules from CT scans. Front Med (Lausanne). 2023;10:1145846. Published 2023 May 19. doi:10.3389/fmed.2023.1145846.

[52] Ottaiano A, Grassi F, Sirica R, et al. Associations between Radiomics and Genomics in Non-Small Cell Lung Cancer Utilizing Computed Tomography and Next-Generation Sequencing: An Exploratory Study. Genes (Basel). 2024;15(6):803. Published 2024 Jun 18. doi:10.3390/genes15060803.

[53] Paez R, Kammer MN, Balar A, et al. Longitudinal lung cancer prediction convolutional neural network model improves the classification of indeterminate pulmonary nodules. Sci Rep. 2023;13(1):6157. Published 2023 Apr 15. doi:10.1038/s41598-023-33098-y.

[54] Park J, Rho MJ, Moon MH. Enhanced deep learning model for precise nodule localization and recurrence risk prediction following curative-intent surgery for lung cancer. PLoS One. 2024;19(7):e0300442. Published 2024 Jul 12. doi:10.1371/journal.pone.0300442.

[55] Prabakaran J, Selvaraj P. Advance IoT Intelligent Healthcare System for Lung Disease Classification Using Ensemble Techniques. Computer Systems Science and Engineering. 2023;46(2):2141–2157. Published 2023 February 9. doi: 10.32604/csse.2023.034210.

[56] Raza R, Zulfiqar F, Khan MO, et al. Lung-EffNet: Lung cancer classification using EfficientNet from CT-scan images. Engineering Applications of Artificial Intelligence. 2023;126:106902. Published 2023 Nov. 10.1016/j.engappai.2023.106902.

[57] Salehjahromi M, Karpinets TV, Sujit SJ, et al. Synthetic PET from CT improves diagnosis and prognosis for lung cancer: Proof of concept. Cell Rep Med. 2024;5(3):101463. doi:10.1016/j.xcrm.2024.101463.

[58] Sousa JV, Matos P, Silva F, Freitas P, Oliveira HP, Pereira T. Single Modality vs. Multimodality: What Works Best for Lung Cancer Screening?. Sensors (Basel). 2023;23(12):5597. Published 2023 Jun 15. doi:10.3390/s23125597.

[59] Tang FH, Fong YW, Yung SH, Wong CK, Tu CL, Chan MT. Radiomics-Clinical AI Model with Probability Weighted Strategy for Prognosis Prediction in Non-Small Cell Lung Cancer. Biomedicines. 2023;11(8):2093. Published 2023 Jul 25. doi:10.3390/biomedicines11082093.

[60] Tonneau M, Phan K, Manem VSK, et al. Generalization optimizing machine learning to improve CT scan radiomics and assess immune checkpoint inhibitors’ response in non-small cell lung cancer: a multicenter cohort study. Front Oncol. 2023;13:1196414. Published 2023 Jul 20. doi:10.3389/fonc.2023.1196414.

[61] Vemula ST, Sreevani M, Rajarajeswari P, Bhargavi K, Tavares MRS, Alankritha S. 2024. Deep Learning Techniques for Lung Cancer Recognition. *Engineering*, Technology & Applied Science Research. 2024;14(4):14916–14922. Published 2024 August. 10.48084/etasr.7510.

[62] Verma S, Magazzù G, Eftekhari N, et al. Cross-attention enables deep learning on limited omics-imaging-clinical data of 130 lung cancer patients. Cell Rep Methods. 2024;4(7):100817. doi:10.1016/j.crmeth.2024.100817.

[63] Wan J, Lin X, Wang Z, et al. Dual-layer spectral detector computed tomography multiparameter machine learning model for prediction of invasive lung adenocarcinoma. Transl Lung Cancer Res. 2025;14(2):385–397. doi:10.21037/tlcr-24-822.

[64] Wang H, Zhu H, Ding L, Yang K. A diagnostic classification of lung nodules using multiple-scale residual network. Sci Rep. 2023;13(1):11322. Published 2023 Jul 13. doi:10.1038/s41598-023-38350-z.

[65] Weiss J, Raghu VK, Bontempi D, et al. Deep learning to estimate lung disease mortality from chest radiographs. Nat Commun. 2023;14(1):2797. Published 2023 May 16. doi:10.1038/s41467-023-37758-5.

[66] Yang S, Lim SH, Hong JH, Park JS, Kim J, Kim HW. Deep learning-based lung cancer risk assessment using chest computed tomography images without pulmonary nodules ≥8 mm. Transl Lung Cancer Res. 2025;14(1):150–162. doi:10.21037/tlcr-24-882.

[67] Yin X, Lu Y, Cui Y, et al. CT-based radiomics-deep learning model predicts occult lymph node metastasis in early-stage lung adenocarcinoma patients: A multicenter study. Chin J Cancer Res. 2025;37(1):12–27. doi:10.21147/j.issn.1000-9604.2025.01.02.

[68] Zhang Z, Zhao Y, Ma YJ, et al. Prediction of STAS in lung adenocarcinoma with nodules ≤ 2 cm using machine learning: a multicenter retrospective study. BMC Cancer. 2025;25(1):417. Published 2025 Mar 7. doi:10.1186/s12885-025-13783-z.

[69] Zheng S, Guo J, Langendijk JA, et al. Survival prediction for stage I-IIIA non-small cell lung cancer using deep learning. Radiother Oncol. 2023;180:109483. doi:10.1016/j.radonc.2023.109483.

[70] Zhou J, Hu B, Feng W, et al. An ensemble deep learning model for risk stratification of invasive lung adenocarcinoma using thin-slice CT. NPJ Digit Med. 2023;6(1):119. Published 2023 Jul 5. doi:10.1038/s41746-023-00866-z.

[71] Zyla J, Marczyk M, Prazuch W, et al. Combining Low-Dose Computer-Tomography-Based Radiomics and Serum Metabolomics for Diagnosis of Malignant Nodules in Participants of Lung Cancer Screening Studies. Biomolecules. 2023;14(1):44. Published 2023 Dec 28. doi:10.3390/biom14010044.

[72] Altalhan M, Algarni A, Turki-Hadj Alouane M. Imbalanced data problem in machine learning: a review. IEEE Access. 2025;13:13686–13699. doi:13.10.1109/ACCESS.2025.3531662.

[73] Ghosh K, Bellinger C, Corizzo R, Branco P, Krawczyk B, Japkowicz N. The class imbalance problem in deep learning. Mach Learn. 2024;113:4845–4901. Published December 28, 2022. doi: 10.1007/s10994-022-06268-8.

[74] Elemento O, Khozin S, Sternberg CN. The use of artificial intelligence for cancer therapeutic decision-making. NEJM AI. 2025;2(5). doi:10.1056/AIra2401164.

[75] Collins GS, Reitsma JB, Altman DG, Moons KG. Transparent reporting of a multivariable prediction model for individual prognosis or diagnosis (TRIPOD): the TRIPOD Statement. BMC Med. 2015;13(1):1. doi:10.1186/s12916-014-0241-z.

[76] Collins GS, Moons KGM, Dhiman P, et al. TRIPOD+AI statement: updated guidance for reporting clinical prediction models that use regression or machine learning methods. BMJ. 2024;385:e078378. Published 2024 Apr 16. doi:10.1136/bmj-2023-078378.

[77] Ong JCL, Chang SYH, William W, et al. Medical ethics of large language models in medicine. NEJM AI. 2024;2(4). doi:10.1056/AIra2400038.

[78] Bianchi F, Queen O, Thakkar N, Sun E, Zou J. Exploring the use of AI authors and reviewers at Agents4Science. Nature Biotechnology. 2025. doi:10.1038/s41587-025-02963-8.

