## SupplementaryMaterials for "Assessing Statistical Practices of Existing Artificial Intelligence (AI) Models for Lung Cancer Detection, Prognosis, and Risk Prediction: A Cross-Sectional Meta-Research Study Supplemented by Human and Large Language Model (LLM)-Directed Quality Appraisal"

**Supplementary Materials Content**

**eMethods 1. (a)** Full keyword query in OpenAlex targeting titles and abstracts and **(b)** prompt for title and abstract screening used in AIScreenR API screened by GPT-4o-mini model.

**eMethods 2**. Human statisticians and clinicians' subjective criteria prior to scoring studies that developed artificial intelligence (AI) tools to predict lung cancer.

**eMethods 3.** Prompts for both GPT-5 Thinking and Gemini 2.5 Pro without assigned roles to evaluate quality score for the 36 studies based on their criteria.

**eMethods 4.** Prompts to assign a clinician (M.D.-level radiologist) and a statistician (Ph.D. level statistician) role for both GPT-5 Thinking and Gemini 2.5 Pro to evaluate quality score for the 36 studies based on their criteria.

**eFigure 1.** Study design and data sources of the 36 included studies.

**eFigure 2. (a)** Bar plot of final model parameters reporting in the 25 studies that used convolutional neural networks (CNNs) as their methods and **(b)** Comparison of available final model parameters versus training data volume with augmentation effect reported in the same 25 studies.

**eFigure 3.** Pairwise Pearson correlation plot on quality scorings by statisticians 1 and 2, clinicians 1 and 2, GPT-5 and Gemini 2.5 Pro without assigned roles averaged over five runs, and GPT-5 and Gemini 2.5 Pro assigned as M.D.-level radiologist averaged over five runs, and GPT-5 and Gemini 2.5 Pro assigned as Ph.D. level statistician averaged over five runs.

**eFigure 4.** Pairwise Pearson correlation plots on quality scorings by Statistician 1 and 2, Clinician 1 and 2, GPT-5 Thinking and Gemini 2.5 Pro without assigned roles averaged over five runs, and GPT-5 Thinking and Gemini 2.5 Pro assigned as M.D.-level radiologist averaged over five runs, and GPT-5 Thinking and Gemini 2.5 Pro assigned as Ph.D. level statistician averaged over five runs.

**eFigure 5.** Pairwise Spearman correlation plots on quality scorings by Statistician 1 and 2, Clinician 1 and 2, GPT-5 Thinking and Gemini 2.5 Pro without assigned roles, and GPT-5 Thinking and Gemini 2.5 Pro assigned as M.D.-level radiologist, and GPT-5 Thinking and Gemini 2.5 Pro assigned as Ph.D. level statistician.

**eResults 1.** Additional description of participant sample sizes of training, sample sizes of testing sets, and validation strategies.

**eResults 2.** Example of the most important criteria for Prompt 2 from a GPT-5 Thinking run without an assigned role.

**eResults 3.** Example of the most important criteria for Prompt 2 from a Gemini 2.5 Pro run without an assigned role.

**eResults 4.** Example of the most important criteria for Prompt 2 from a GPT-5 Thinking run assigned as a Ph.D. level statistician.

**eResults 5.** Example of the most important criteria for Prompt 2 from a Gemini 2.5 Pro run assigned as a Ph.D. level statistician.

**eResults 6.** Example of the most important criteria for Prompt 2 from a GPT-5 Thinking run assigned as a M.D.-level radiologist.

**eResults 7.** Example of the most important criteria for Prompt 2 from a Gemini 2.5 Pro run assigned as a M.D.-level radiologist.

**eResults 8.** Overlap of evaluation criteria from CHARMS guideline and both GPT-5 and Gemini 2.5 Pro without role assignment.

**eTable 1.** CHARMS 2014 relevant items to extract from individual studies in a systematic review of prediction models, evaluated for our meta-research study assessing artificial intelligence (AI) for lung cancer risk prediction.

**eTable 2.** Study characteristics of the 36 included studies stratified by outcome endpoints, with five studies double-counted. There were 14 studies that reported detection, 18 studies reported prognosis, and 9 studies reported risk prediction.

**eTable 3.** Pre-trained model architectures reported in the 25 studies that used convolutional neural networks (CNNs) in their methods.

**eTable 4.** Methods for handling imbalanced outcome classes during training among the 36 included studies.

**eTable 5.** Quality scorings by Statistician 1 and 2, Clinician 1 and 2, GPT-5 Thinking and Gemini 2.5 Pro without assigned roles, and GPT-5 Thinking and Gemini 2.5 Pro assigned as M.D.-level radiologist, and GPT-5 Thinking and Gemini 2.5 Pro assigned as Ph.D. level statistician..

**eMethods 1.**

**(a)** Full keyword query in OpenAlex targeting titles and abstracts

("AI" OR "artificial intelligence" OR "machine learning" OR "ML" OR "deep learning" OR "DL") AND ("predict" OR "prediction" OR "predictions") AND ("CT" OR "computed tomography" OR "MRI" OR "magnetic resonance imaging" OR "x-ray" OR "x ray" OR "PET" OR "positron emission tomography") AND ("detection" OR "screening" OR "risk") AND "cancer"

**(b)** Prompt for title and abstract screening for the AIScreenR API screened by GPT-4o-mini model

The prompt used was: “Evaluate the following study based on the selection criteria for a review of the design and evaluation metrics of research studies that use artificial intelligence to identify or predict lung cancer from medical imaging. The study must be about the creation of some sort of new artificial intelligence tool that is using medical images to screen for, identify, or predict the risk of lung cancer by analyzing images.”

**eMethods 2**. Human statisticians and clinicians' subjective criteria prior to scoring studies that developed artificial intelligence (AI) tools to predict lung cancer.

**Statisticians:** Prioritized clarity and rigor of reporting, sound methodological design, overall reproducibility, model generalizability, transparency of model explanation, and coherent organization.

**Clinicians:** Prioritized the appropriateness of the study cohort and follow-up, clarity of outcome definitions, model performance, whether results support the conclusions, and applicability to the patient groups (and subpopulations) in which the model would be used.

**eMethods 3.** Prompts for both GPT-5 and Gemini 2.5 Pro without assigned roles to evaluate quality scores for the 36 studies based on their criteria.

[Prompt 1] *Use the re-run button to run 10 times in a single chat conversation.

We have a project that aims to evaluate published studies that develop artificial intelligence (AI) models using imaging data for lung cancer detection, prognosis, and risk prediction.

Can you please design a comprehensive set of evaluation criteria to assess published research papers from January 1, 2023, to June 30, 2025, on AI models proposed for lung cancer detection, prognosis, and prediction from imaging data. The framework must evaluate a paper’s scientific rigor, training, testing, and validation of models, clinical relevance, and potential for generalizability. You may consider available statistical, clinical, and regulatory guidelines. List your criteria.

[Prompt 2] *Model’s complete list (total of 10 lists) should be in a Word document attached to this prompt.

Here is a large list of possible criteria to consider for assessing published research studies from January 1, 2023, to June 30, 2025, on AI models for lung cancer detection, prognosis and prediction from imaging data. Please choose the most important criteria from this list that evaluate the scientific rigor, training, testing, and validation of models, clinical relevance, and potential for generalizability of the work. Based on your chosen criteria, score each study on a 1–10 scale, where 10 = excellent adherence to the criteria and 1 = very poor adherence.

For each study, provide:

The score (1–10).

A brief justification that explains why the study received this score, referring directly to the evaluation criteria.

We have a total of 36 studies, and we will upload their PDFs one by one.

**eMethods 4.** Prompts to assign a M.D.-level radiologist and a Ph.D.-level statistician role for both GPT-5 and Gemini 2.5 Pro to evaluate quality scores for the 36 studies based on their criteria.

[Prompt 1] *Use the re-run button to run 10 times in a single chat conversation.

We have a project that aims to evaluate published studies that develop artificial intelligence (AI) models using imaging data for lung cancer detection, prognosis, and risk prediction.

Assume you are an M.D.-level and board-certified radiologist who screens for lung cancer in the United States, can you please design a comprehensive set of evaluation criteria to assess published research papers from January 1, 2023, to June 30, 2025, on AI models proposed for lung cancer detection, prognosis, and prediction from imaging data. The framework must evaluate a paper’s scientific rigor, training, testing, and validation of models, clinical relevance, and potential for generalizability. You may consider available statistical, clinical, and regulatory guidelines. List your criteria.

[Prompt 2] *Model’s complete list (total of 10 lists) should be in a Word document attached to this prompt.

Here is a large list of possible criteria to consider for assessing published research studies from January 1, 2023, to June 30, 2025, on AI models for lung cancer detection, prognosis and prediction from imaging data. Please choose the most important criteria from this list that evaluate the scientific rigor, training, testing, and validation of models, clinical relevance, and potential for generalizability of the work. Based on your chosen criteria, score each study on a 1–10 scale, where 10 = excellent adherence to the criteria and 1 = very poor adherence.

For each study, provide:

The score (1–10).

A brief justification that explains why the study received this score, referring directly to the evaluation criteria.

We have a total of 36 studies, and we will upload their PDFs one by one.

[Prompt 1] *Use the re-run button to run 10 times in a single chat conversation.

We have a project that aims to evaluate published studies that develop artificial intelligence (AI) models using imaging data for lung cancer detection, prognosis, and risk prediction.

Assume you are a Ph.D.-level statistician with 10 years of working experience in the United States, can you please design a comprehensive set of evaluation criteria to assess published research papers from January 1, 2023, to June 30, 2025, on AI models proposed for lung cancer detection, prognosis, and prediction from imaging data. The framework must evaluate a paper’s scientific rigor, training, testing, and validation of models, clinical relevance, and potential for generalizability. You may consider available statistical, clinical, and regulatory guidelines. List your criteria.

[Prompt 2] *Model’s complete list (total of 10 lists) should be in a Word document attached to this prompt.

Here is a large list of possible criteria to consider for assessing published research studies from January 1, 2023, to June 30, 2025, on AI models for lung cancer detection, prognosis and prediction from imaging data. Please choose the most important criteria from this list that evaluate the scientific rigor, training, testing, and validation of models, clinical relevance, and potential for generalizability of the work. Based on your chosen criteria, score each study on a 1–10 scale, where 10 = excellent adherence to the criteria and 1 = very poor adherence.

For each study, provide:

The score (1–10).

A brief justification that explains why the study received this score, referring directly to the evaluation criteria.

We have a total of 36 studies, and we will upload their PDFs one by one.

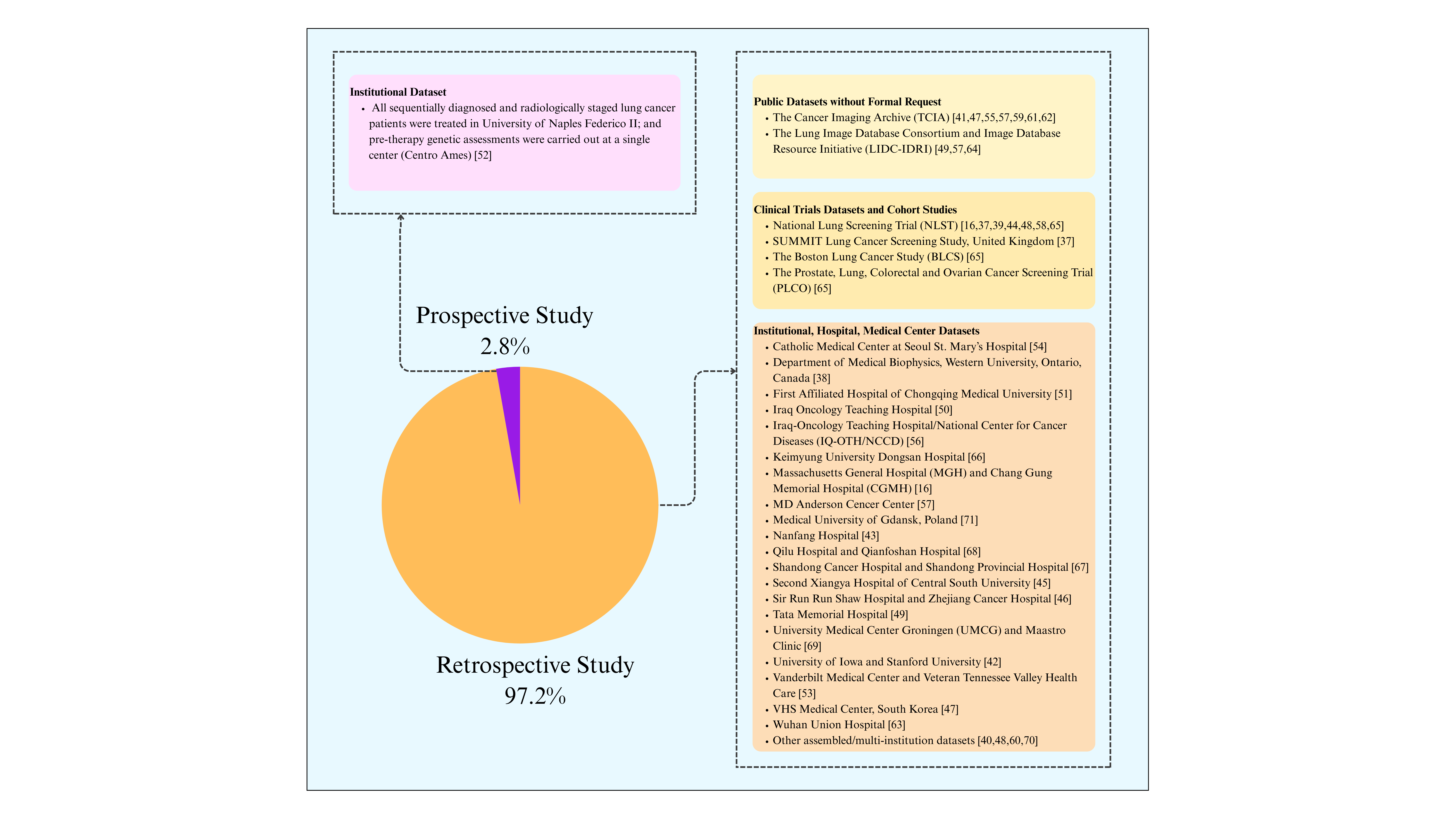

**eFigure 1.** Study design and data sources of the 36 included studies.

**(a)**

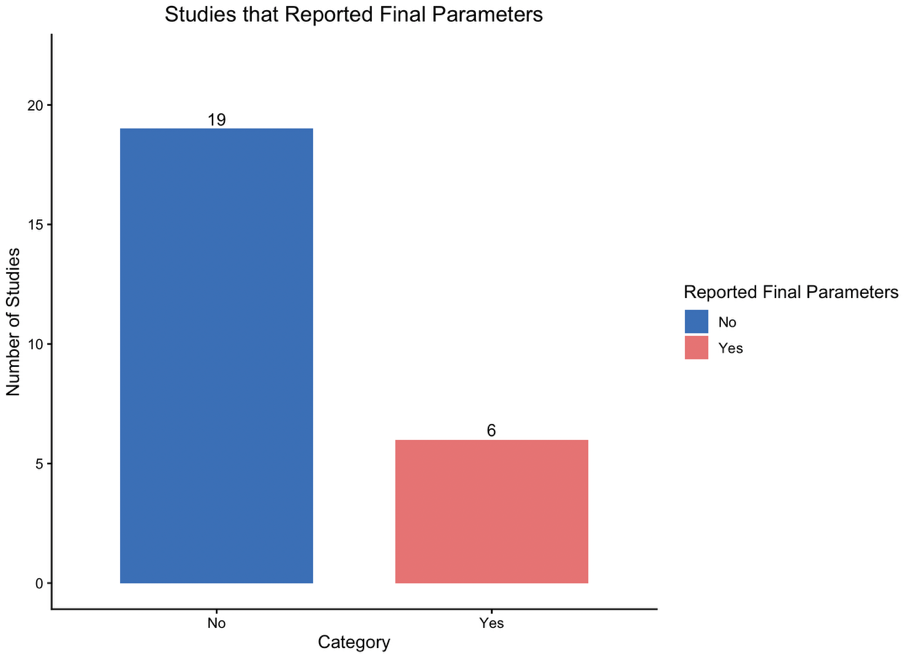

**(b)**

Note: Arrows indicate the increase in training data size due to data augmentation for Prabakaran et al. [1] and Raza et al. [2].

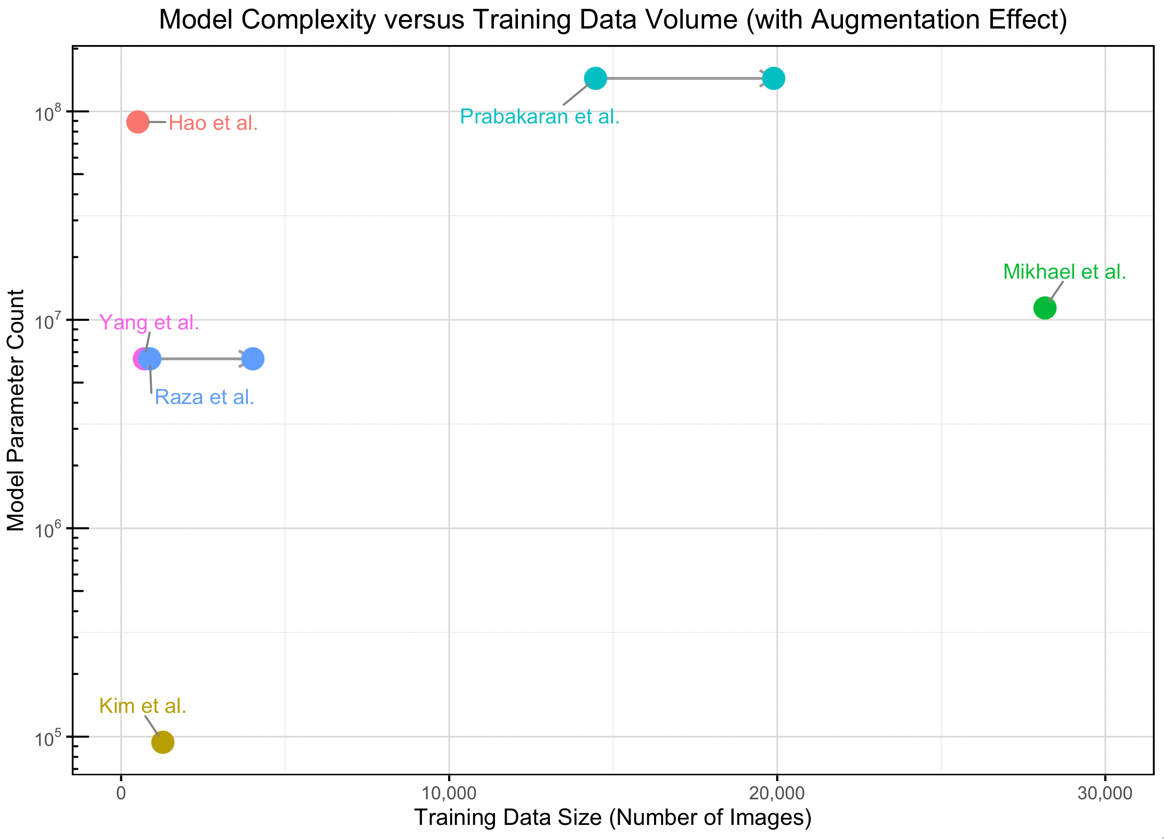

**eFigure 2. (a)** Bar plot of final model parameters reporting in the 25 studies that used convolutional neural networks (CNNs) as their methods; **(b)** Comparison of available final model parameters versus training data volume with augmentation effect reported in the same 25 studies.

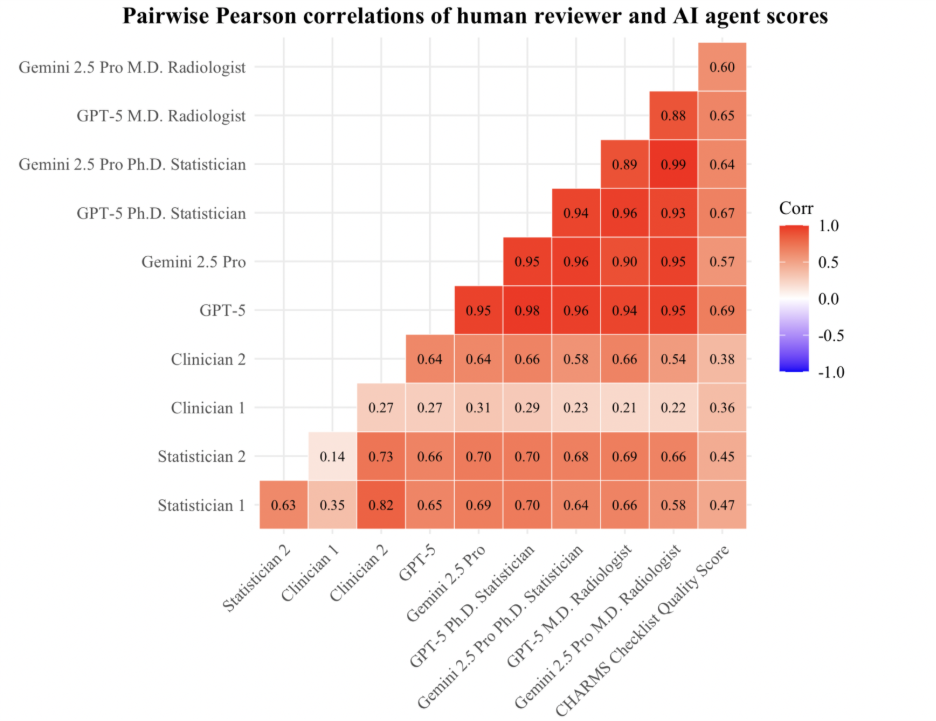

**eFigure 3.** Pairwise Pearson correlation plot on quality scorings by statisticians 1 and 2, clinicians 1 and 2, GPT-5 and Gemini 2.5 Pro without assigned roles averaged over five runs, and GPT-5 and Gemini 2.5 Pro assigned as M.D.-level radiologist averaged over five runs, and GPT-5 and Gemini 2.5 Pro assigned as Ph.D. level statistician averaged over five runs.

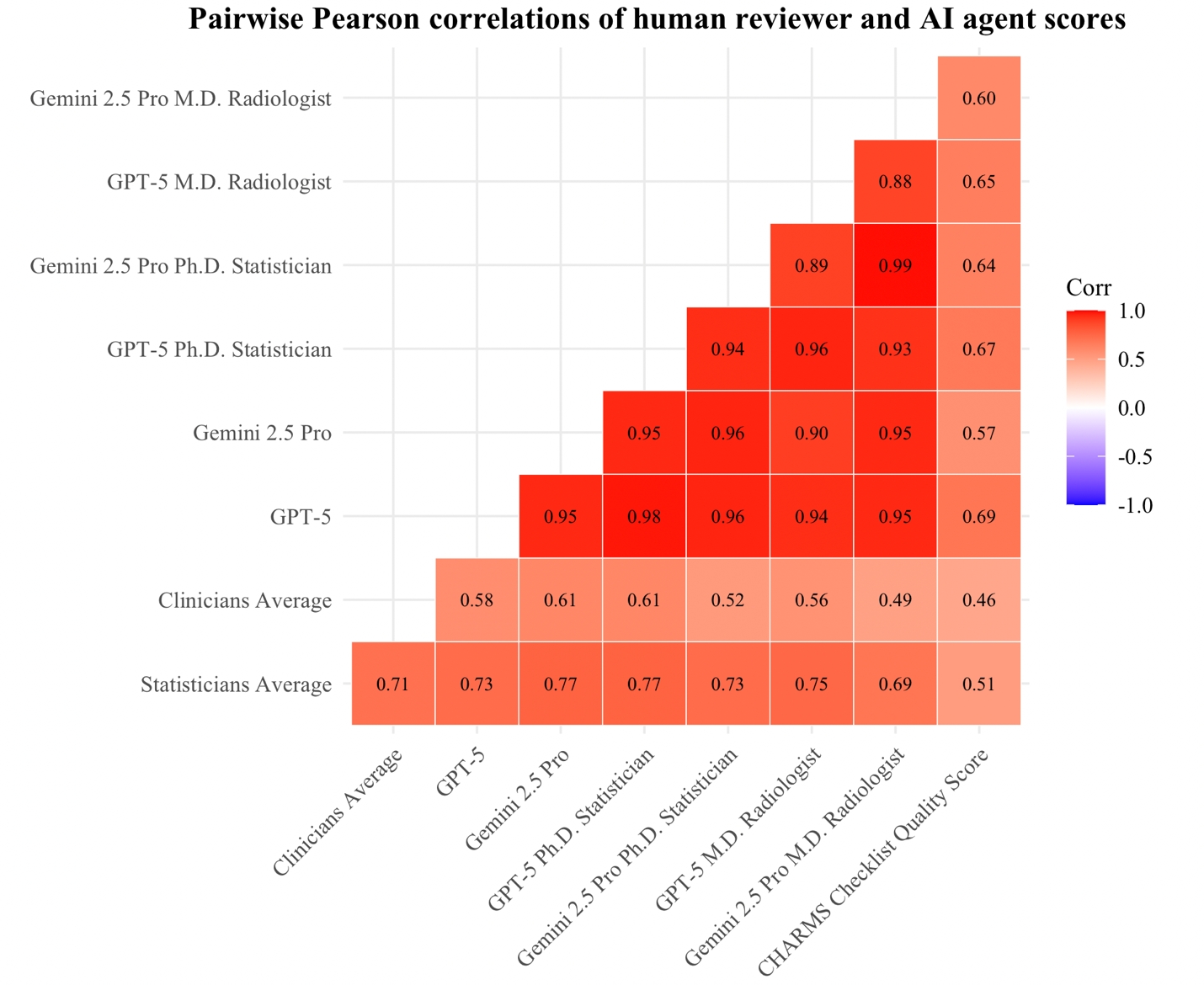
**eFigure 4.** Pairwise Pearson correlation plots on quality scorings by average of statisticians 1 and 2, average of clinicians 1 and 2, GPT-5 and Gemini 2.5 Pro without assigned roles average over five runs, and GPT-5 and Gemini 2.5 Pro assigned as M.D.-level radiologist over five runs, and GPT-5 and Gemini 2.5 Pro assigned as Ph.D.-level statistician over five runs.

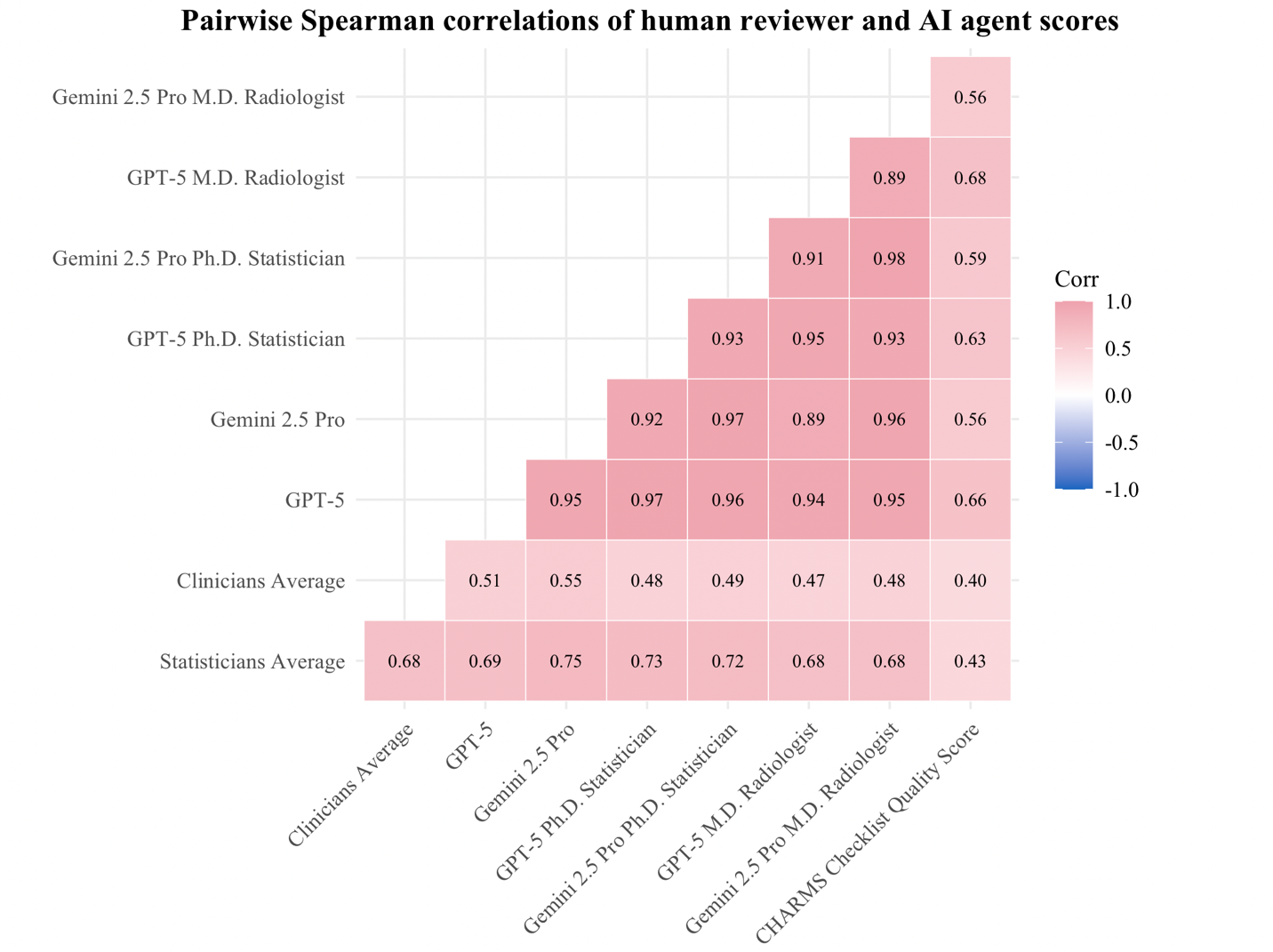

**eFigure 5.** Pairwise Spearman correlation plots on quality scorings by average of statisticians 1 and 2, average of clinicians 1 and 2, GPT-5 Thinking and Gemini 2.5 Pro without assigned roles, and GPT-5 and Gemini 2.5 Pro assigned as M.D.-level radiologist, and GPT-5 and Gemini 2.5 Pro assigned as Ph.D.-level statistician.

**eResults 1.** Additional description of participant sample sizes of training, sample sizes of testing sets, and validation strategies.

Training dataset sizes of participants varied widely: two studies trained models on fewer than 100 samples; 16 studies trained on 100 to 500 samples; 9 studies trained on 500 to 1,000 samples; and 3 studies trained on 1,000 or more samples. Six studies did not report its training sample size. Internal hold-out test set was used in 27 of the 36 included studies (75%), with 1 study using two internal test sets [3]. Among these 27 studies, 12 studies validated their models on internal test sets smaller than 100 participant samples; 6 studies used sets of 100 to 500 participant samples; 4 studies used sets larger than 500 participant samples; and 6 study did not report the size of its internal test set. There were 8 studies that only used cross-validation instead of a fixed internal hold-out test.

External validation was reported in 14 of the 36 included studies (39%), with 4 studies that used multiple external test sets [4,5,6,7]. External test sample sizes varied, with 6 studies using fewer than 100 participant samples, 7 studies using between 10 and 200 participant samples, 3 studies using between 200 and 500 participant samples, 1 study using between 500 and 2,000 participant samples, and 3 studies using more than 2,000 participant samples.

**eResults 2.** Example of the most important criteria for Prompt 2 from a GPT-5 Thinking run without an assigned role.

### **Core scoring rubric (10 points total)**

1. Clinical task & study design (0–1)
    Clear intended use (screening/triage/detection/diagnosis/segmentation/prognosis/risk), target users/setting, proper design (retrospective vs prospective; reader or impact study if claims imply it), and pre-specified endpoints/analysis plan.
2. Cohort provenance & representativeness (0–1)
    Multi-site/vendor/time span reported; inclusion/exclusion and participant flow diagram; demographics and disease spectrum reflect intended population; sample size/events justified.
3. Ground truth quality (0–1)
    Appropriate reference standard (e.g., pathology, adjudicated longitudinal outcome), rater expertise/number, adjudication and inter-rater agreement; clear outcome windows and censoring/competing risks (when relevant).
4. Splits & leakage control (0–1)
    Strict patient-level separation of train/val/test; no near-duplicate/series leakage; transforms fit on train only; site/temporal hold-out if feasible.
5. Model development transparency (0–1)
    Architecture, pretraining source/domain, inputs, loss, hyperparameters, augmentation, early stopping/regularization, and seeds/versions reported; imbalance strategy stated (e.g., focal loss/weights/sampling) with rationale.
6. Internal validation soundness (0–1)
    Proper CV or bootstrap (nested if tuning), thresholds locked using validation—not test; ablations/sensitivity analyses for key choices (e.g., with/without clinical covariates, segmentation dependency).
7. Metrics, uncertainty & calibration (0–1)
    Task-appropriate primary metrics with 95% CIs; detection uses lesion-/case-level FROC or FP/scan; prognosis uses C-index/time-AUC and Brier; classification reports AUPRC when imbalanced; calibration curves/slope/intercept (or ECE) and uncertainty estimates or at least bootstrap CIs.
8. External validation & transportability (0–1)
    At least one independent test differing by site/time or vendor; report performance deltas and per-site/vendor results; basic shift/robustness checks (e.g., slice thickness, reconstruction kernel, dose).
9. Clinical relevance & human comparison (0–1)
    Strong baselines (radiologists, Lung-RADS/Brock/SoC) and/or reader study; decision-analytic utility (net benefit/threshold justification), workload/read-time impact, clear operating point tied to clinical consequences.
10. Transparency, reporting standards & sharing (0–1)
     Adherence to fit-for-purpose checklists (TRIPOD-AI/CLAIM/STARD-AI; CONSORT-AI/DECIDE-AI if applicable), data/code/model card availability or justified access path; explicit limitations, failure-mode review, and generalizability statement.

#### **How I’ll score (anchors)**

- 1 = Fully meets item (clear evidence; solid execution).
- 0.5 = Partially meets (some gaps/ambiguity).
- 0 = Not met (missing/inadequate or undermines validity).

##### **Automatic downgrades (apply once, in addition to item scores)**

- Proven leakage or test-set peeking: cap total at **≤3**.
- No external validation but making broad deployment/generalization claims: cap at **≤6**.
- Inappropriate/insufficient ground truth for the claim (e.g., diagnostic claims without credible reference standard): cap at **≤5**.

**eResults 3.** Example of the most important criteria for Prompt 2 from a Gemini 2.5 Pro run without an assigned role.

**Evaluation Criteria Framework**

- I have selected the following 10 core criteria, organized into four key domains, which will form the basis for the scoring.

##### **1. 🔬 Scientific Rigor and Study Design**

- 1. Clarity of Clinical Objective & Cohort:
- The study must have a clearly defined clinical question and specify the target population with transparent **inclusion and exclusion criteria**.
- 2. Quality of Reference Standard:
- The "ground truth" used for training and testing must be robust and clearly described, with **histopathological confirmation** being the gold standard.
- 3. Adherence to Reporting Guidelines:
- The paper should follow established guidelines like **TRIPOD or STARD-AI** to ensure transparency and completeness.

##### **2. ✅ Model Validation and Reproducibility**

- 4. Rigorous Data Splitting:
- There must be a strict, **patient-level split** of data into independent training, validation, and testing sets to prevent data leakage.
- **5. External Validation:** This is a critical criterion. The model's performance must be tested on at least one
- **completely independent external dataset** from a different institution, population, or time period.
- 6. Appropriate Metrics & Code Availability:
- The study must report task-appropriate metrics (e.g., AUC, C-index) with **95% confidence intervals** and, ideally, make its
- **code publicly available**.

##### **3. 🩺 Clinical Relevance and Utility**

- 7. Comparison to a Clinical Benchmark:
- The model’s performance must be compared against a relevant standard, such as the performance of **human experts (radiologists)** or existing risk models.
- 8. Interpretability and Workflow Integration:
- The study should provide insight into the model's decisions using **explainability techniques** (e.g., saliency maps) and discuss how the tool could be integrated into a clinical workflow.

##### **4. 🌍 Generalizability and Robustness**

- 9. Data Diversity:
- The training and validation data should be sourced from **multiple institutions** and reflect demographic and clinical diversity to support generalizability.
- 10. Subgroup and Error Analysis:
- The study should analyze model performance across relevant **patient subgroups** and investigate its failure cases to understand its limitations.

#### **Scoring Framework (1–10 Scale)**

- **10 (Excellent):** The study meets all 10 criteria. It is likely a prospective, multi-center study with robust external validation on diverse cohorts, transparently reported with publicly available code, and a clear path to clinical utility.
- **8–9 (Very Good):** The study is methodologically strong and meets most criteria, including the critical requirement of external validation. It may have minor weaknesses, such as a less diverse external cohort or limited discussion of workflow integration.
- **6–7 (Good):** A solid study with a key limitation. Typically, this study has strong internal validation but **lacks external validation**, or it uses a single-center dataset, limiting its generalizability. The core methodology is sound, but its real-world performance is unproven.
- **4–5 (Fair):** The study has significant methodological flaws. It may lack a separate test set, have unclear data splitting procedures, or fail to compare against a relevant clinical benchmark. The conclusions are likely not fully supported.
- **2–3 (Poor):** The study has multiple major flaws, such as a high risk of data leakage, use of inappropriate metrics, and a lack of detail needed for reproducibility. The results are not reliable.
- **1 (Very Poor):** The study is fundamentally flawed across most criteria and provides no credible evidence to support its claims.

**eResults 4.** Example of the most important criteria for Prompt 2 from a GPT-5 Thinking run assigned as a Ph.D. level statistician.

### **Core domains & weights (100 pts total)**

1. Clinical question & intended use (8)

- Task, users, setting, and actionability are explicit (e.g., LDCT screening triage vs. diagnostic aid).
- Outcomes are guideline-relevant and time-anchored (e.g., Lung-RADS categories, OS/PFS).

2. Cohort design & reference standard (12)

- Transparent cohort assembly (in/exclusion, timeframe, prevalence), flow diagram.
- Ground truth fit to task (pathology/longitudinal outcome/consensus) with blinding & inter-rater reliability.

3. Imaging protocols & preprocessing transparency (8)

- Scanner/vendor, dose, slice thickness, kernel; preprocessing/harmonization fully specified.
- Segmentation approach & QC; radiomics steps IBSI-consistent when applicable.

4. Data partitions & leakage control (10)

- Patient-level splits; no ROI/test leakage; thresholds tuned only on validation; locked test set(s).
- Prefer temporal/site/device external holdout.

5. Sample size, events & imbalance/missingness (6)

- Justified sample size/EPV; prevalence reported; principled class-imbalance & missing-data handling.

6. Model development & training rigor (10)

- Architecture/features, hyperparameter search, regularization/early stopping, augmentations documented.
- Ablations & strong baselines (e.g., Brock/PLCOm2012, radiologist, clinical-only).

7. Metrics & statistical analysis (12)

- Task-appropriate primary endpoints with 95% CIs (e.g., FROC/JAFROC for detection; C-index/time-AUC for survival).
- Proper paired tests/bootstraps; per-patient (and per-lesion when relevant) reporting.

8. Calibration & clinical utility (8)

- Reliability plots, calibration slope/intercept (or Brier/IBS).
- Decision-curve analysis/net benefit at clinically chosen thresholds; confusion matrices at prespecified cutoffs.

9. External validation & transportability (12)

- At least one site/temporal/device-external test with performance deltas; subgroup reporting by site/vendor/protocol.
- Stress/shift tests (dose, kernel, noise) or test–retest where feasible.

10. Fairness & subgroup performance (5)

- Sex/age/smoking/race/ethnicity (when available) subgroup metrics with uncertainty; mitigation or candid limits.

11. Transparency, reproducibility & reporting standards (7)

- TRIPOD-AI/CLAIM/STARD-AI adherence; code/weights/configs or credible access statement; model/data cards; exact splits.
- PROBAST-AI/QUADAS-2 risk-of-bias/applicability addressed.

12. Regulatory & lifecycle readiness (2)

- Stated intended SaMD-style use, monitoring/update plan (e.g., PCCP concept), privacy/security; workflow integration notes.

#### **Converting to a 1–10 study score**

Compute the 0–100 total, then map:

- 91–100 → 10 (Outstanding)
- 81–90 → 9
- 71–80 → 8
- 61–70 → 7
- 51–60 → 6
- 41–50 → 5
- 31–40 → 4
- 21–30 → 3
- 11–20 → 2
- 0–10 → 1

#### **What to record for each study (tight template)**

**Overall score (1–10):** X/10
 **Why:** 1–2 sentences tying to the biggest wins/gaps.

**Domain ticks (brief):**

1. Clinical use ( /8):
2. Cohort & reference ( /12):
3. Imaging & preprocessing ( /8):
4. Splits & leakage ( /10):
5. Size/imbalance/missing ( /6):
6. Model training rigor ( /10):
7. Metrics & stats ( /12):
8. Calibration & utility ( /8):
9. External validation ( /12):
10. Fairness ( /5):
11. Transparency/reporting ( /7):
12. Regulatory/lifecycle ( /2):

**One-line justification example:**
 “Strong multi-site external validation with decision-curve analysis (+), clear leakage control (+), but limited subgroup/fairness reporting and no calibration (−), hence 8/10.”

#### **Notes on *what matters most* when scoring**

- Heaviest signal of generalizability: true site/device/temporal **external validation** with clear deltas and subgroup breakdowns.
- Heaviest signal of clinical relevance: **calibration + decision-curve** at policy-relevant thresholds (not AUC alone).
- Biggest validity risks: **leakage**, unreported **prevalence/imbalance**, and lack of **reference-standard blinding**.

**eResults 5.** Example of the most important criteria for Prompt 2 from a Gemini 2.5 Pro run assigned as a Ph.D. level statistician.

##### **## Core Evaluation Criteria 🎯**

I will assess each paper against these five core criteria, which synthesize the most important points from the provided list.

1. **Study Design & Scientific Rigor:** This evaluates the foundation of the study.
   a. **Key elements:** A clearly defined clinical objective (detection, prognosis, or risk prediction) ; a strong study design (prospective and/or multi-center designs are superior to single-center retrospective ones) ; and the use of an appropriate, independent reference standard (e.g., histopathology for diagnosis).
2. **External Validation & Generalizability:** This is the most crucial test of a model's real-world potential.
   a. **Key elements:** The model's performance **must be validated on at least one completely independent external cohort** from a different institution, population, or time period. A study's score is fundamentally limited without this. Performance on diverse data (e.g., different scanner types) is also assessed.
3. **Data Partitioning & Technical Integrity:** This assesses the technical soundness of the model's development.
   a. **Key elements:** A strict, **patient-level separation** of data into independent training, validation, and testing sets to prevent data leakage. The methods must be described in enough detail for reproducibility, with the public availability of code and/or model weights being a major strength.
4. **Statistical Analysis & Performance Reporting:** This evaluates the robustness of the results.
   a. **Key elements:** Use of appropriate performance metrics for the task (e.g., AUC for detection, C-index for prognosis). Critically, all key metrics **must be reported with 95% confidence intervals** to quantify uncertainty. For risk models, an assessment of **model calibration** is essential.
5. **Clinical Relevance & Impact:** This assesses the practical value and safety of the model.
   a. **Key elements:** A fair comparison to a relevant benchmark (e.g., radiologists, existing clinical models). Analysis of performance in clinical subgroups and an investigation of the model's failures (error analysis) are necessary to understand its limitations. The inclusion of a clinical utility assessment, like Decision Curve Analysis, is a sign of a high-impact study.

##### **## Scoring Rubric (1–10 Scale) ⚖️**

Each study will be assigned a single score based on its overall adherence to the criteria above.

- **10 (Excellent):** The study is methodologically exceptional. It features a prospective or strong multi-center design, rigorous external validation on diverse data, impeccable and transparent methods (e.g., code is available), robust statistical analysis (including CIs and calibration), and a clear, compelling demonstration of clinical relevance.
- **8–9 (Very Good):** A strong study with robust findings. It has most of the elements of a top-tier paper, such as multi-center data and proper external validation, but may have minor limitations, like limited code availability or a less thorough analysis of clinical utility.
- **6–7 (Good):** A solid scientific contribution. The study is well-designed but may be limited to a single institution or have less extensive external validation. The statistical analysis is sound, but may lack deeper investigation into calibration or subgroup performance. The model shows promise but requires more evidence for generalizability.
- **4–5 (Fair):** The study has a valid concept but contains significant methodological weaknesses that limit the credibility of its conclusions. This often includes studies with **no external validation**, relying only on an internal test set. The generalizability of the findings is highly questionable.
- **1–3 (Poor):** The study has critical, disqualifying flaws. This may include data leakage between training and testing sets, a lack of any independent test set, grossly inappropriate statistical metrics, or a failure to describe methods sufficiently. The results are considered untrustworthy.

**eResults 6.** Example of the most important criteria for Prompt 2 from a GPT-5 Thinking run assigned as a M.D.-level radiologist.

### **Scoring rubric (100 → 1–10)**

1. **Clinical question & intended use (U.S. workflow fit)** – 8
    Is the task (LDCT screening triage, nodule detection/characterization, prognosis/risk) explicit, mapped to a real decision (e.g., Lung-RADS change, biopsy, follow-up interval), with the target users and setting clear?
2. **Cohort construction & representativeness** – 10
    Consecutive/registry sampling, inclusion/exclusion appropriate, demographics & smoking history reported, spectrum reflects intended population (screening vs symptomatic), IRB/consent noted.
3. **Ground truth quality** – 10
    Task-appropriate reference: pathology or longitudinal outcome for malignancy; consensus thoracic-rad reads for detection; blinded labeling; inter-rater agreement reported.
4. **Data provenance & splits (leakage control)** – 10
    Patient-level train/val/test separation; temporal or site-held-out test; no preprocessing or tuning on test; clear counts per split.
5. **External validation (transportability)** – 12
    Independent site/time/vendor; thresholds frozen from development; performance retained with CIs; (prospective/silent-mode earns full credit).
6. **Performance metrics matched to task + uncertainty** – 12
    Detection: FROC and sensitivity at fixed FP/scan; Classification: AUROC + AUPRC with operating points; Prognosis: C-index/time-AUC; **All** with 95% CIs and proper paired tests.
7. **Calibration & decision impact** – 8
    Calibration plots/Brier; thresholds justified by clinical costs or guideline mapping; decision-curve analysis or net benefit; confusion matrices at chosen operating points.
8. **Comparators to standard of care** – 8
    Strong, fair baselines (e.g., Lung-RADS, Brock/PLCOm2012, radiologists with MRMC where applicable) and statistically sound comparisons.
9. **Robustness, subgroup equity & shift analysis** – 10
    Stratified by site/scanner/sex/age/race/ethnicity/smoking; sensitivity to dose, kernel, slice thickness, motion; missing-data strategy; disparity gaps quantified (with CIs) and addressed.
10. **Model development transparency** – 6
     Architecture/feature set, hyperparameters, augmentation, class-imbalance handling, initialization/pretraining, segmentation QA (if used) are reproducible.
11. **Reader study & workflow integration (if claimed)** – 6
     MRMC/crossover design, time-to-read, impact on sensitivity/specificity and variability; clear assist mode (triage/second-read), alert burden.
12. **Reporting, reproducibility & regulatory signals** – 10
     CLAIM/TRIPOD-AI/STARD-AI/DECIDE-AI (as appropriate) addressed; code/weights or credible reproduction path; intended-use statement and SaMD mindset (monitoring, change control).

**Gatekeeper rule:** If **no external validation**, cap the total at **70/100** unless the study is explicitly methodological.

#### **Quick scoring template (use this per paper)**

- **Overall score (1–10):** X.X
- **Why:** 1) Clinical fit: …; 2) Cohort/GT: …; 3) Splits/external: …; 4) Metrics/CIs & calibration: …; 5) Comparators: …; 6) Robustness/equity: …; 7) Repro/regulatory: … (1–2 concise lines each).

#### **What I’ll look for (translation of the rubric into checks)**

- **Intended use & decision point** are explicit and align with **U.S. LDCT/Lung-RADS** when relevant.
- **Leakage-safe design** with **held-out external** data; thresholds **locked** before testing.
- **Task-appropriate metrics** + **CIs** and **calibration**; **decision-curve/net-benefit** ties numbers to patient care.
- **Fair head-to-head** against **Lung-RADS / Brock / radiologists**.
- **Subgroup/shift analyses** to prove generalizability; articulate **failure modes**.
- **Reproducible** methods & **reporting checklists** satisfied; signs of **regulatory readiness** (intended use, monitoring).

**eResults 7.** Example of the most important criteria for Prompt 2 from a Gemini 2.5 Pro run assigned as a M.D.-level radiologist.

##### **Essential Evaluation Criteria**

I will assess each paper against these five non-negotiable domains, which represent the absolute cornerstones of high-quality, clinically relevant AI research.

1. **Clinical Problem and Ground Truth**
   a. **Addresses a specific, significant clinical need** (e.g., reducing false positives, predicting treatment response).
   b. **Uses a "gold standard" ground truth**: Histopathology for diagnosis or hard clinical endpoints (like Overall Survival) for prognosis.
2. **Data Sourcing and Generalizability**
   a. **Multi-Institutional Data**: The model must be trained and/or tested on data from multiple, diverse institutions to ensure it's not overfit to a single center's protocols or patient population.
   b. **External Validation**: The model's performance must be verified on a completely independent, unseen external test set from a different institution, time period, or geography. This is the single most critical test of generalizability.
3. **Methodological and Statistical Rigor**
   a. **Patient-Level Data Splitting**: The paper must confirm that data was split into training, validation, and test sets at the patient level to prevent data leakage.
   b. **Appropriate Metrics with Confidence Intervals**: The study must report clinically relevant metrics (e.g., AUC-ROC, C-index) accompanied by 95% confidence intervals to quantify statistical uncertainty.
   c. **Model Calibration**: The study must assess if the model's predicted probabilities are accurate (e.g., with a calibration plot).
4. **Clinical Benchmarking**
   a. **Comparison to Standard of Care**: The model's performance must be directly benchmarked against the relevant clinical standard. This means a comparison against board-certified radiologists for diagnostic tasks or against the TNM staging system for prognostic tasks.
5. **Transparency and Reproducibility**
   a. **Failure Analysis**: The paper should include an analysis of the model's failure modes (false positives and false negatives) to understand its limitations.
   b. **Adherence to Reporting Guidelines**: The work should follow established guidelines like TRIPOD or STARD to ensure completeness and clarity.

##### **Scoring Rubric (1–10 Scale)**

I will assign a single score to each paper based on its overall adherence to the essential criteria above.

- **10 (Excellent):** Meets or exceeds all essential criteria. The study features robust, multi-institutional external validation, is benchmarked against clinicians, demonstrates high statistical rigor (including calibration), and is presented with exceptional transparency. The results are highly credible and show clear potential for clinical translation.
- **8–9 (Very Good):** A methodologically strong paper that meets most key criteria. It likely uses multi-institutional data and has rigorous internal validation but may have a slightly less robust external validation cohort or minor gaps in reporting (e.g., missing a failure analysis). Generalizability is highly likely.
- **6–7 (Good):** A solid study with some noteworthy limitations. For example, it might be a single-institution study, which fundamentally limits claims of generalizability. While methodologically sound, the lack of data diversity or external validation means the model's real-world performance is unproven.
- **4–5 (Average):** The study is promising but has significant methodological weaknesses. It may lack a held-out test set, use a weak ground truth (e.g., imaging follow-up instead of pathology) , or fail to report confidence intervals. The results should be viewed as preliminary and exploratory.
- **2–3 (Poor):** Contains at least one major methodological flaw. This could include clear data leakage (e.g., not splitting by patient), failure to benchmark against any clinical standard, or using inappropriate metrics. The study's conclusions are not well-supported.
- **1 (Very Poor):** A critically flawed study with multiple major errors. The methodology is unsound, the reporting is incomplete, and the results are not reliable or reproducible.

**eResults 8.** Overlap of evaluation criteria from CHARMS guideline and both GPT-5 and Gemini 2.5 Pro without role assignment.

Here we describe the overlap between evaluation criteria from CHARMS and both GPT-5 and Gemini 2.5 Pro without role assignment (example of criteria output in eResults 2-3). Like the CHARMS guideline, both GPT-5 and Gemini’s criteria required transparent description of data and cohort selection, clear outcome definitions, explicit model building and validation procedures, model interpretation, and generalizability. The GPT-5 rubric further noted risks specific to modern imaging analysis using AI—tight leakage controls, patient-level splits, pretraining disclosure, and mandatory external test sets—while also including factors for clinical importance such as utility of the AI model, failure-mode review, and data/code/model card availability. Gemini covers similar content in four domains (1. Scientific rigor and study design, 2. Model validation and reproducibility, 3. Clinical relevance and utility 4. Generalizability and robustness), however, its guidance is more narrative and less granular compared to the structured criteria from GPT-5 and CHARMS. Overall, CHARMS provides the foundational reporting framework for prediction models, while the GPT-5 and Gemini criteria adapt and extend these expectations for studies developing modern AI imaging models for lung cancer outcomes.

**eTable 1.** CHARMS 2014 relevant items to extract from individual studies in a systematic review of prediction models, evaluated for our meta-research study assessing artificial intelligence (AI) for lung cancer risk prediction.

| **Domain** | **Key items** |
| --- | --- |
| **SOURCE OF DATA** | Source of data (e.g., cohort, case-control, randomized trial participants, or registry data) |
| **PARTICIPANTS** | Participant eligibility and recruitment method (e.g., consecutive participants, location, number of centers, setting, inclusion and exclusion criteria) |
|  | Participant description |
|  | Details of treatments received, if relevant |
|  | Study dates |
| **OUTCOME(S) TO BE PREDICTED** | Definition and method for measurement of outcome |
|  | Was the same outcome definition (and method for measurement) used in all patients? |
|  | Type of outcome (e.g., single or combined endpoints) |
|  | ~~Was the outcome assessed without knowledge of the candidate predictors (i.e., blinded)?~~ |
|  | ~~Were candidate predictors part of the outcome (e.g., in panel or consensus diagnosis)?~~ |
|  | Time of outcome occurrence or summary of duration of follow-up |
| **CANDIDATE PREDICTORS**  **(OR INDEX TESTS)** | Number and type of predictors (e.g., demographics, patient history, physical examination, additional testing, disease characteristics) |
|  | Definition and method for measurement of candidate predictors |
|  | Timing of predictor measurement (e.g., at patient presentation, at diagnosis, at treatment initiation) |
|  | ~~Were predictors assessed blinded for outcome, and for each other (if relevant)?~~ |
|  | Handling of predictors in the modelling (e.g., continuous, linear, non-linear transformations or categorised) |
| **SAMPLE SIZE** | Number of participants and number of outcomes/events |
|  | Number of outcomes/events in relation to the number of candidate predictors (Events Per Variable) |
| **MISSING DATA (Only applicable to studies that had or commented on missing data)** | Number of participants with any missing value (include predictors and outcomes) |
|  | Number of participants with missing data for each predictor |
|  | Handling of missing data (e.g., complete-case analysis, imputation, or other methods) |
| **MODEL DEVELOPMENT** | Modelling method (e.g., logistic, survival, neural network, or machine learning techniques) |
|  | Modelling assumptions satisfied |
|  | Method for selection of predictors **for inclusion** in multivariable modelling (e.g., all candidate predictors, pre-selection based on unadjusted association with the outcome) |
|  | Method for selection of predictors **during multivariable modelling** (e.g., full model approach, backward or forward selection) and criteria used (e.g., p-value, Akaike Information Criterion) |
|  | Shrinkage of predictor weights or regression coefficients (e.g., no shrinkage, uniform shrinkage, penalized estimation) |
| **MODEL PERFORMANCE** | Calibration (calibration plot, calibration slope, Hosmer-Lemeshow test) and Discrimination (C-statistic, D-statistic, log-rank) measures with confidence intervals |
|  | Classification measures (e.g., sensitivity, specificity, predictive values, net reclassification improvement) and whether a-priori cut points were used |
| **MODEL EVALUATION** | Method used for testing model performance: development dataset only (random split of data, resampling methods e.g. bootstrap or cross-validation, none) or separate external validation (e.g. temporal, geographical, different setting, different investigators) |
|  | In case of poor validation, whether model was adjusted or updated (e.g., intercept recalibrated, predictor effects adjusted, or new predictors added) |
| **RESULTS** | Final and other multivariable models (e.g., basic, extended, simplified) presented, including predictor weights or regression coefficients, intercept, baseline survival, model performance measures (with standard errors or confidence intervals) |
|  | Any alternative presentation of the final prediction models, e.g., sum score, nomogram, score chart, predictions for specific risk subgroups with performance |
|  | Comparison of the distribution of predictors (including missing data) for development and validation datasets |
| **INTERPRETATION AND DISCUSSION** | Interpretation of presented models (confirmatory, i.e., model useful for practice versus exploratory, i.e., more research needed) |
|  | Comparison with other studies, discussion of generalizability, strengths and limitations. |

**eTable 2.** Study characteristics of the 36 included studies stratified by outcome endpoints, with five studies double-counted. There were 14 studies that reported detection, 18 studies reported prognosis, and 9 studies reported risk prediction.

|  | Detection | Prognosis | Risk prediction |  | Detection | Prognosis | Risk prediction |
| --- | --- | --- | --- | --- | --- | --- | --- |
|  | Y N | Y N | Y N |  | Counts | Counts | Counts |
| Input Data  Demographics  Clinical | 3 11  3 11 | 8 10  9 9 | 2 7  2 7 | **Reported Uncertainty Quantification**  None  Evaluation Metrics  Others | 7  6  0 | 4  10  4 | 1  7  1 |
| Data/Code Availability  Data  Code | 8 6  3 11 | 6 12  2 16 | 6 3  4 5 | **Source Population**  Europe  Asia  North America | 1  7  8 | 2  8  9 | 0  3  7 |
| Subgroup Evaluation | 2 12 | 4 14 | 4 5 | **Training Size**  < 100  100-500  500-1000  > 1000  Not Reported | 0  5  2  1  6 | 1  10  5  1  1 | 1  4  2  2  0 |
| Test Set Category  Internal  External | 14 0  2 12 | 18 0  8 10 | 8 1  6 3 | **Internal Test Size**  < 100  100-500  > 500  Cross Validation  Not Reported | 4  3  1  0  6 | 9  2  1  6  1 | 1  2  3  2  0 |
| Evaluation Metric  AUC  Sensitivity  Accuracy  Specificity  F1 Score | Counts  10  10  11  6  7 | Counts  15  11  10  9  4 | Counts  8  3  3  3  2 | **External Test Size**  < 100  100-200  200-500  500-2000  > 2000 | 0  3  1  1  0 | 3  4  1  0  0 | 3  3  2  1  3 |
| Precision  Calibration plot  C-Index  PPV  NPV  DCA  Confusion  matrix  FROC | 4  1  2  3  3  1  2  2 | 4  4  2  1  1  4  2  0 | 0  1  2  1  1  0  0  0 | **Algorithm Type**  CNN  SVM  RF  LR  GBM  Ensemble  DT  Autoencoder  ViT  RNN  GLM  cGAN | 10  2  3  2  2  0  0  1  0  0  0  1 | 12  4  3  5  5  2  2  0  1  0  1  0 | 6  2  2  1  3  0  0  2  0  1  0  1 |

Algorithm type abbreviations are Convolutional Neural Network (CNN), Decision Tree (DT), conditional Generative Adversarial Network (cGAN), Gradient Boosting Machine (GBM), Generalized Linear Model (GLM), Logistic Regression (LR), Random Forest (RF), Recurrent Neural Network (RNN), Support Vector Machine (SVM), Vision Transformer (ViT).Evaluation metric abbreviations are area under the curve (AUC), concordance index (C-index), decision curve analysis (DCA), positive predictive value (PPV), negative predictive value (NPV), free-response receiver operating characteristic (FROC).

**eTable 3.** Pre-trained model architectures reported in the 25 studies that used convolutional neural networks (CNNs) in their methods.

Abbreviations: CNN for convolutional neural network; LCP for Lung Cancer Prediction; VGG for visual geometry group

**List of pre-trained model architecture**

Did not report use of pre-trained models (9 studies).

**For model building:**

2D ConvNexr-B — explicit note on pretraining on ImageNet-22K [8].

DenseNet-201 [9].

Efficient Net-B0 [9].

EfficientNet-B0-B4 — explicit note on pretraining on ImageNet [2].

Inception-ResNet-v2 [1].

Inception-v4 [7].

3D Inception [11].

3D LCP-CNN [12].

MobileNet v2 [10].

3D ResNet [11].

3D ResNet-18 [4,13,14].

3D ResNet-34 [7,15,16].

ResNet-50 [1].

SE-ResNet-18 [10].

3D VGG [11].

VGG-16 [13,17,18].

VGG-19 [1,13,18].

**For segmentation or preprocessing:**

3D CT and PET U-Net [42].

2D Dense U-Net [10].

**eTable 4.** Methods for handling imbalanced outcome classes during training among the 36 included studies.

**Methods of handling imbalanced outcome classes during training**

Did not report using any method to address imbalanced data (22 studies).

Explicitly balanced data for each outcome class [13,16,20,21].

Minority target data augmentation [1,2,22].

Synthetic minority over-sampling technique (SMOTE) [6,23].

Sigmoid focal cross-entropy in convolutional neural network (CNN) models [24].

Logit-adjusted cross-entropy to make multi-class survival classifier robust to imbalanced time-to-death bins [25].

Class weighted binary cross-entropy and focal loss in CNN models [26]

Sampling weights for imbalanced classes [27].

Class weights usage [14].

**eTable 5.** Quality scorings by Statistician 1 and 2, Clinician 1 and 2, GPT-5 and Gemini 2.5 Pro without assigned roles, and GPT-5 and Gemini 2.5 Pro assigned as M.D.-level radiologist, and GPT-5 and Gemini 2.5 Pro assigned as Ph.D.-level statistician.

Please find quality scorings in table Supplementary_eTable5.docx

**References**

[1] Prabakaran J, Selvaraj P. Advance IoT Intelligent Healthcare System for Lung Disease Classification Using Ensemble Techniques. *Computer Systems Science and Engineering*. 2023;46(2):2141-2157. Published 2023 February 9. doi: 10.32604/csse. 2023.034210.

[2] Raza R, Zulfiqar F, Khan MO, et al. Lung-EffNet: Lung cancer classification using EfficientNet from CT-scan images. Engineering Applications of Artificial Intelligence. 2023;126:106902. Published 2023 Nov. <https://doi.org/10.1016/j.engappai.2023.106902.>

[3] Hao P, Yu Y, Huang CT, et al. Advancing EGFR mutation subtypes prediction in NSCLC by combining 3D pretrained ConvNeXt, radiomics, and clinical features. *Front Oncol*. 2024;14:1464555. Published 2024 Nov 15. doi:10.3389/fonc.2024.1464555.

[4] Mikhael PG, Wohlwend J, Yala A, et al. Sybil: A Validated Deep Learning Model to Predict Future Lung Cancer Risk From a Single Low-Dose Chest Computed Tomography. *J Clin Oncol*. 2023;41(12):2191-2200. doi:10.1200/JCO.22.01345.

[5] Salehjahromi M, Karpinets TV, Sujit SJ, et al. Synthetic PET from CT improves diagnosis and prognosis for lung cancer: Proof of concept. *Cell Rep Med*. 2024;5(3):101463. doi:10.1016/j.xcrm.2024.101463.

[6] Verma S, Magazzù G, Eftekhari N, et al. Cross-attention enables deep learning on limited omics-imaging-clinical data of 130 lung cancer patients. *Cell Rep Methods*. 2024;4(7):100817. doi:10.1016/j.crmeth.2024.100817.

[7] Weiss J, Raghu VK, Bontempi D, et al. Deep learning to estimate lung disease mortality from chest radiographs. *Nat Commun*. 2023;14(1):2797. Published 2023 May 16. doi:10.1038/s41467-023-37758-5.

[8] Hao P, Yu Y, Huang CT, et al. Advancing EGFR mutation subtypes prediction in NSCLC by combining 3D pretrained ConvNeXt, radiomics, and clinical features. *Front Oncol*. 2024;14:1464555. Published 2024 Nov 15. doi:10.3389/fonc.2024.1464555.

[9] Hu D, Li X, Lin C, Wu Y, Jiang H. Deep Learning to Predict the Cell Proliferation and Prognosis of Non-Small Cell Lung Cancer Based on FDG-PET/CT Images. *Diagnostics (Basel)*. 2023;13(19):3107. Published 2023 Sep 30. doi:10.3390/diagnostics13193107.

[10] Yang S, Lim SH, Hong JH, Park JS, Kim J, Kim HW. Deep learning-based lung cancer risk assessment using chest computed tomography images without pulmonary nodules ≥8 mm. *Transl Lung Cancer Res*. 2025;14(1):150-162. doi:10.21037/tlcr-24-882.

[11] Zhou J, Hu B, Feng W, et al. An ensemble deep learning model for risk stratification of invasive lung adenocarcinoma using thin-slice CT. *NPJ Digit Med*. 2023;6(1):119. Published 2023 Jul 5. doi:10.1038/s41746-023-00866-z.

[12] Paez R, Kammer MN, Balar A, et al. Longitudinal lung cancer prediction convolutional neural network model improves the classification of indeterminate pulmonary nodules. *Sci Rep*. 2023;13(1):6157. Published 2023 Apr 15. doi:10.1038/s41598-023-33098-y.

[13] Kim G, Park YM, Yoon HJ, Choi JH. A multi-kernel and multi-scale learning based deep ensemble model for predicting recurrence of non-small cell lung cancer. *PeerJ Comput Sci*. 2023;9:e1311. Published 2023 May 2. doi:10.7717/peerj-cs.1311.

[14] Sousa JV, Matos P, Silva F, Freitas P, Oliveira HP, Pereira T. Single Modality vs. Multimodality: What Works Best for Lung Cancer Screening?. *Sensors (Basel)*. 2023;23(12):5597. Published 2023 Jun 15. doi:10.3390/s23125597.

[15] Wang H, Zhu H, Ding L, Yang K. A diagnostic classification of lung nodules using multiple-scale residual network. *Sci Rep*. 2023;13(1):11322. Published 2023 Jul 13. doi:10.1038/s41598-023-38350-z.

[16] Yin X, Lu Y, Cui Y, et al. CT-based radiomics-deep learning model predicts occult lymph node metastasis in early-stage lung adenocarcinoma patients: A multicenter study. *Chin J Cancer Res*. 2025;37(1):12-27. doi:10.21147/j.issn.1000-9604.2025.01.02.

[17] Tonneau M, Phan K, Manem VSK, et al. Generalization optimizing machine learning to improve CT scan radiomics and assess immune checkpoint inhibitors' response in non-small cell lung cancer: a multicenter cohort study. *Front Oncol*. 2023;13:1196414. Published 2023 Jul 20. doi:10.3389/fonc.2023.1196414.

[18] Vemula ST, Sreevani M, Rajarajeswari P, Bhargavi K, Tavares MRS, Alankritha S. 2024. Deep Learning Techniques for Lung Cancer Recognition. *Engineering, Technology & Applied Science Research*. 2024;14(4):14916-149221. Published 2024 August. DOI:https://doi.org/10.48084/etasr.7510.

[19] Gainey JC, He Y, Zhu R, et al. Predictive power of deep-learning segmentation based prognostication model in non-small cell lung cancer. *Front Oncol*. 2023;13:868471. Published 2023 Apr 4. doi:10.3389/fonc.2023.868471.

[20] Aslani S, Alluri P, Gudmundsson E, et al. Enhancing cancer prediction in challenging screen-detected incident lung nodules using time-series deep learning. *Comput Med Imaging Graph*. 2024;116:102399. doi:10.1016/j.compmedimag.2024.102399.

[21] Tang FH, Fong YW, Yung SH, Wong CK, Tu CL, Chan MT. Radiomics-Clinical AI Model with Probability Weighted Strategy for Prognosis Prediction in Non-Small Cell Lung Cancer. *Biomedicines*. 2023;11(8):2093. Published 2023 Jul 25. doi:10.3390/biomedicines11082093.

[22] Maijeddah UI, Yusuf SA, Abdullhai M, Hassan IH. A hybrid transfer learning model with optimized SVM using honey badger optimization algorithm for multi-class lung cancer classification. *Science World Journal*. 2025;19(4):977-986. Published 2025 February 14. doi: 10.4314/swj.v19i4.10.

[23] Ebrahimpour L, Després P, Manem VSK. Differential Radiomics-Based Signature Predicts Lung Cancer Risk Accounting for Imaging Parameters in NLST Cohort. *Cancer Med*. 2024;13(20):e70359. doi:10.1002/cam4.70359.

[24] Eldho KJ, Nithyanandh S. (2024) Lung Cancer Detection and Severity Analysis with a 3D Deep Learning CNN Model Using CT-DICOM Clinical Dataset. *Indian Journal of Science and Technology*. 17(10): 899-910. <https://doi.org/10.17485/IJST/v17i10.3085.>

[25] Hermoza R, Nascimento JC, Carneiro G. Weakly-supervised preclinical tumor localization associated with survival prediction from lung cancer screening Chest X-ray images. *Comput Med Imaging Graph*. 2024;115:102395. doi:10.1016/j.compmedimag.2024.102395.

[26] Zheng S, Guo J, Langendijk JA, et al. Survival prediction for stage I-IIIA non-small cell lung cancer using deep learning. *Radiother Oncol*. 2023;180:109483. doi:10.1016/j.radonc.2023.109483.

[27] Zyla J, Marczyk M, Prazuch W, et al. Combining Low-Dose Computer-Tomography-Based Radiomics and Serum Metabolomics for Diagnosis of Malignant Nodules in Participants of Lung Cancer Screening Studies. *Biomolecules*. 2023;14(1):44. Published 2023 Dec 28. doi:10.3390/biom14010044.
