## Supplementary_eTable5 for "Assessing Statistical Practices of Existing Artificial Intelligence (AI) Models for Lung Cancer Detection, Prognosis, and Risk Prediction: A Cross-Sectional Meta-Research Study Supplemented by Human and Large Language Model (LLM)-Directed Quality Appraisal"

**eTable 5.** Quality scorings by statisticians 1 and 2, clinicians 1 and 2, GPT-5 and Gemini 2.5 Pro without assigned roles, GPT-5 and Gemini 2.5 Pro assigned as an M.D.-level and board-certified radiologist who screens for lung cancer in the United States, and GPT-5 and Gemini 2.5 Pro assigned as a Ph.D.-level statistician with 10 years of working experience in the United States.

| Authors | Statistician 1 | Statistician 2 | Clinician 1 | Clinician 2 | GPT-5  (Average over 5 runs) | Gemini 2.5 Pro (Average over 5 runs) | GPT-5 Ph.D. Statistician in the US (Average over 5 runs) | Gemini 2.5 Pro Ph.D. Statistician in the US (Average over 5 runs) | GPT-5 M.D. Radiologist in the US (Average over 5 runs) | Gemini 2.5 Pro M.D. Radiologist in the US (Average over 5 runs) | CHARMS Checklist Quality Score |
| --- | --- | --- | --- | --- | --- | --- | --- | --- | --- | --- | --- |
| Aslani  et al. [37]  05/20/24 | 8 | 6.8 | 7 | 7 | 7.22 | 8.2 | 7.28 | 8.4 | 7.32 | 8.2 | 8.3 |
| Christie et al. [38]  05/03/25 | 9 | 6.3 | 6 | 8 | 7.70 | 8.8 | 7.56 | 8.8 | 7.58 | 8.8 | 9.3 |
| Ebrahimpour et al. [39]  10/28/24 | 8 | 4.8 | 8 | 8 | 6.10 | 3.8 | 6.28 | 4.4 | 6.14 | 3.4 | 9.1 |
| Eldho  et al. [40]  02/27/24 | 5 | 2.5 | 6 | 5 | 2.96 | 1.0 | 3.04 | 1.6 | 2.96 | 1.2 | 6.9 |
| Fanizzi  et al. [41]  11/23/23 | 6 | 7.0 | 7 | 7 | 5.60 | 5.2 | 5.30 | 5.0 | 5.12 | 4.8 | 8.6 |
| Gainey  et al. [42]  04/03/23 | 8 | 5.8 | 9 | 8 | 6.42 | 7.4 | 6.26 | 7.2 | 5.62 | 7.6 | 8.3 |
| Hao et al. [43]  11/14/24 | 9 | 4.8 | 9 | 8 | 6.14 | 5.8 | 5.86 | 5.4 | 5.62 | 4.6 | 8.6 |
| Hermoza et al. [44]  07/01/24 | 9 | 4.8 | 7 | 9 | 5.86 | 6.2 | 5.56 | 5.2 | 5.38 | 4.6 | 6.6 |

| Authors | Statistician 1 | Statistician 2 | Clinician 1 | Clinician 2 | GPT-5  (Average over 5 runs) | Gemini 2.5 Pro (Average over 5 runs) | GPT-5 Ph.D. Statistician (Average over 5 runs) | Gemini 2.5 Pro Ph.D. Statistician (Average over 5 runs) | GPT-5 M.D. Radiologist (Average over 5 runs) | Gemini 2.5 Pro M.D. Radiologist (Average over 5 runs | CHARMS Checklist Quality Score |
| --- | --- | --- | --- | --- | --- | --- | --- | --- | --- | --- | --- |
| Hu et al. [45]  09/30/23 | 6 | 3.8 | 8 | 7 | 5.58 | 3.2 | 5.16 | 4.2 | 5.14 | 4.2 | 7.9 |
| Huang  et al. [46]  03/19/24 | 8 | 8.0 | 7 | 8 | 7.80 | 8.4 | 7.30 | 9.2 | 7.08 | 9.6 | 9.0 |
| Kim et al. [47]  05/02/23 | 8 | 7.5 | 7 | 8 | 6.14 | 5.6 | 6.00 | 5.2 | 5.56 | 5.0 | 7.9 |
| Liu et al. [48]  06/05/24 | 8 | 5.3 | 8 | 8 | 6.82 | 8.6 | 6.24 | 8.0 | 5.76 | 7.6 | 9.0 |
| Mahajan et al. [49]  02/08/25 | 8 | 5.0 | 8 | 7 | 6.02 | 7.2 | 5.94 | 6.2 | 5.52 | 5.8 | 6.6 |
| Maijeddah et al. [50]  02/14/25 | 7 | 3.5 | 7 | 6 | 3.34 | 1.4 | 3.40 | 2.8 | 2.76 | 1.2 | 8.6 |
| Mikhael et al. [16]  01/12/23 | 10 | 9.5 | 8 | 9 | 8.74 | 10.0 | 8.40 | 10.0 | 8.42 | 10.0 | 10.0 |
| Mu et al. [51]  05/18/23 | 7 | 4.3 | 9 | 7 | 6.20 | 6.8 | 5.66 | 6.0 | 5.72 | 6.4 | 8.3 |
| Ottaiano et al. [52]  06/18/24 | 6 | 3.8 | 8 | 6 | 4.76 | 3.2 | 4.52 | 3.6 | 4.20 | 3.0 | 8.6 |
| Paez  et al. [53]  04/15/23 | 6 | 5.3 | 7 | 7 | 5.84 | 4.8 | 5.46 | 4.6 | 5.66 | 4.4 | 8.6 |
| Park  et al. [54]  07/12/24 | 7 | 5.0 | 8 | 7 | 3.84 | 2.4 | 4.12 | 2.2 | 3.52 | 1.6 | 7.9 |

| Authors | Statistician 1 | Statistician 2 | Clinician 1 | Clinician 2 | GPT-5  (Average over 5 runs) | Gemini 2.5 Pro (Average over 5 runs) | GPT-5 Ph.D. Statistician (Average over 5 runs) | Gemini 2.5 Pro Ph.D. Statistician (Average over 5 runs) | GPT-5 M.D. Radiologist (Average over 5 runs) | Gemini 2.5 Pro M.D. Radiologist (Average over 5 runs) | CHARMS Checklist Quality Score |
| --- | --- | --- | --- | --- | --- | --- | --- | --- | --- | --- | --- |
| Prabakaran et al. [55]  02/09/23 | 7 | 3.5 | 7 | 6 | 3.40 | 1.8 | 3.66 | 2.4 | 2.94 | 1.6 | 7.2 |
| Raza  et al. [56]  09/03/23 | 6 | 3.5 | 7 | 7 | 3.86 | 1.8 | 3.68 | 2.0 | 4.34 | 1.2 | 6.9 |
| Salehjahromi  et al. [57]  03/19/24 | 9 | 9.3 | 9 | 9 | 7.62 | 10.0 | 7.26 | 9.8 | 6.48 | 10.0 | 8.6 |
| Sousa  et al. [58]  06/15/23 | 7 | 4.5 | 8 | 6 | 5.42 | 5.2 | 5.12 | 5.2 | 5.24 | 4.6 | 8.3 |
| Tang  et al. [59]  07/25/23 | 8 | 3.5 | 7 | 7 | 5.70 | 5.6 | 5.64 | 5.8 | 5.68 | 5.8 | 7.8 |
| Tonneau et al. [60]  07/19/23 | 7 | 4.8 | 8 | 7 | 7.42 | 9.0 | 7.12 | 9.0 | 6.10 | 9.6 | 8.3 |
| Vemula et al. [61]  05/07/24 | 5 | 3.3 | 6 | 6 | 2.90 | 1.2 | 3.14 | 2.2 | 3.78 | 1.6 | 6.2 |
| Verma  et al. [62]  07/15/24 | 9 | 8.0 | 7 | 8 | 7.40 | 8.4 | 6.84 | 9.4 | 6.42 | 8.8 | 7.6 |
| Wan et al. [63]  02/27/25 | 7 | 5.5 | 8 | 7 | 6.70 | 6.6 | 6.30 | 6.8 | 5.78 | 6.0 | 8.6 |

| Authors | Statistician 1 | Statistician 2 | Clinician 1 | Clinician 2 | GPT-5  (Average over 5 runs) | Gemini 2.5 Pro (Average over 5 runs) | GPT-5 Ph.D. Statistician (Average over 5 runs) | Gemini 2.5 Pro Ph.D. Statistician (Average over 5 runs) | GPT-5 M.D. Radiologist (Average over 5 runs) | Gemini 2.5 Pro M.D. Radiologist (Average over 5 runs) | CHARMS Checklist Quality Score |
| --- | --- | --- | --- | --- | --- | --- | --- | --- | --- | --- | --- |
| Wang et al. [64]  07/13/23 | 7 | 6.8 | 8 | 7 | 4.08 | 5.0 | 4.84 | 2.8 | 4.94 | 2.0 | 6.9 |
| Weiss  et al. [65]  04/27/23 | 9 | 8.8 | 7 | 9 | 8.20 | 9.2 | 7.02 | 10.0 | 7.60 | 9.6 | 9.7 |
| Yang  et al. [66]  01/24/25 | 9 | 5.5 | 9 | 8 | 7.08 | 7.8 | 6.74 | 7.4 | 6.74 | 5.8 | 9.7 |
| Yin et al. [67]  01/24/25 | 7 | 4.3 | 8 | 6 | 7.98 | 8.2 | 7.08 | 8.8 | 6.28 | 9.0 | 9.0 |
| Zhang et al. [68]  03/07/25 | 8 | 5.0 | 8 | 7 | 7.52 | 8.4 | 7.02 | 8.8 | 7.14 | 9.2 | 9.7 |
| Zheng  et al. [69]  01/20/23 | 6 | 5.3 | 6 | 7 | 7.24 | 7.4 | 6.50 | 7.8 | 6.02 | 7.6 | 7.9 |
| Zhou  et al. [70]  07/05/23 | 7 | 6.0 | 8 | 7 | 7.50 | 8.4 | 7.00 | 9.2 | 7.20 | 9.4 | 9.0 |
| Zyla et al. [71]  12/28/23 | 7 | 3.5 | 7 | 7 | 5.84 | 5.4 | 5.20 | 4.4 | 5.26 | 3.4 | 7.6 |
